# Short-term postprandial glucose monitoring reveals stable traits from noisy free-living meals

**DOI:** 10.64898/2026.09.14.26362991

**Authors:** M. Toumi, M. Salathé

## Abstract

Postprandial glucose responses differ greatly between meals, which complicates the identification of individual glucose response traits in free-living conditions. We analysed 50,463 free-living meals from 992 adults without diagnosed diabetes using a multivariate mixed-effects machine-learning framework and evaluated transfer to 4,524 standardized meals excluded from training. This model separated the population-shared from participant-specific patterns across four joint glucose response outcomes. Within-individual variability dominated free-living responses, yet the between-individual variation was concentrated in two dimensions. The dominant axis represented an overall tendency toward larger glucose excursions. The second axis reflected carbohydrate responsiveness with lower late glucose elevations. Dominant-axis scores were reproducible across non-overlapping meal subsets and associated with held-out standardized-meal responses, beyond age, sex, BMI, and glucose mean and variability. Dominant-axis scores estimated from 3 days of monitoring approximated those estimated from 14 days. These findings support short-term monitoring to measure glucose response individuality and identify a candidate coordinate for stratification.

## Introduction

Postprandial glucose responses (PPGR) provide repeated measures of metabolic regulation in everyday life. Even in individuals without diagnosed diabetes, large excursions are recognized as an early marker of impaired glucose control [1]. However, meals with similar nutritional compositions can produce largely different excursions across individuals, both under controlled conditions and in everyday settings [2], [3], which limits the usefulness of generalized dietary guidance. This heterogeneity is partly explained by individual factors such as gut microbiome composition, physical activity, anthropometrics and lifestyle factors [4]. These findings suggest that PPGR heterogeneity may reflect biological differences between individuals.

Free-living glycemic heterogeneity may originate from two complementary sources of biological variation. First, it is well known that individual glycemic responses are primarily influenced by the eating context, including meal composition, prior dietary intake, physical activity [5], and diurnal physiology [6]. These factors result in complex individual-specific response patterns and obscure the shared response structure across individuals. Second, individuals may also differ in physiological aspects of glucose homeostasis, including insulin secretion and sensitivity [7]. These differences are partly obscured under free-living conditions, as suggested by standardized meal-challenge and dynamic glucose-insulin modeling studies [8], [9].

These explanations lead to different approaches to precision nutrition. If glucose responses depend only on the meal context, dietary guidance could be restricted to the context characteristics and applied across individuals, regardless of their personal glucose-response traits. If responses are also driven by physiological differences, dietary guidance would need to account for variation between individuals. Progress along this path has been limited because metabolic individuality is difficult to isolate and identify under free-living conditions. Variation in meal and glycemic contexts contributes to high within-individual fluctuations, which makes glucose responses an imprecise measure of the underlying physiology.

Various personalized-nutrition frameworks were designed to model and predict individual postprandial responses. These models use person-level covariates such as anthropometrics and microbiome composition as personalizing features, and are evaluated for predictive performance in previously unseen individuals [1], [4], [10]. These approaches have shown that PPGR can be predicted with useful accuracy, but are optimized for predictive performance across populations. These models generally predict each meal response separately, without explicitly separating differences between individuals from context-dependent variations.

Meanwhile, repeated standardized carbohydrate challenges have demonstrated reproducible individual response differences linked to metabolic physiology [11]. CGM-based approaches such as the “glucotypes” framework [12] have identified phenotypes from glucose level dynamics, suggesting that recurrent patterns of glucose variability can identify distinct glucose control profiles. Complementary functional analyses of postprandial trajectories from free-living evening meals have separated participant- and meal-level modes of variation and found that dominant person-level differences largely reflect overall glucose level [13]. However, these approaches decompose the observed CGM trajectory variations and provide less insight into the structure of persistent response traits after adjustment for context. Furthermore, Sugimoto et al. [14] showed that mean glucose, glucose variance, and autocorrelation explain more than 80% of interindividual variation in CGM-derived measures. However, their work was based on overall glucose dynamics rather than responses to free-living meals.

We used a phenotype-first approach: we modelled the between-individual variation in glucose response as structured deviations from a population-shared response function. These individual-specific deviations captured uniform shifts in glycemic response and differences in how glucose responses scaled with macronutrient intakes. We fitted a multivariate mixed-effects machine-learning model with 50,463 free-living meals collected in 992 adults without diagnosed diabetes, and investigated if persistent glucose response traits could be identified from routine monitoring alone. We found that individual responses were organized along a low-dimensional glycemic phenotype space. Its first axis reflected overall glycemic elevation: an individual’s overall tendency towards larger or smaller glucose responses. The second axis combined carbohydrate responsiveness with lower late glucose elevations. When we conditioned the macronutrient slopes on the response intercepts, carbohydrate responsiveness remained structured along a distinct carbohydrate-versus-fiber axis. These results support the recovery of such traits from routine self-monitoring, shifting the modelling goal from single-meal predictions to metabolic phenotyping.

## Materials and Methods

### Cohort and Data collection

We analysed data from the *Food & You* study [15], a digital nutrition cohort of 1,014 adults in Switzerland who completed free-living monitoring of diet, interstitial glucose levels, and lifestyle variables. All participants followed the same protocol with 14 days of monitoring in cohort B (N=870) and 28 days in cohort C (N=144 women), which also collected menstrual-cycle data. This cohort included adults living in Switzerland without diagnosed diabetes. Exclusion criteria included pregnancy and use of glucose-lowering medications. Participants wore FreeStyle Libre 2 sensors that recorded interstitial glucose every 15 minutes throughout the monitoring period, yielding approximately 1.47 million glucose readings in total. Dietary intakes were logged in real time via the MyFoodRepo smartphone application, through photo-based entries, text entries and barcode scans. Entries were reviewed by trained human annotators following a standardized annotation protocol, and nutritional content was derived using the MyFoodRepo food composition database, which integrates branded-product label information and standardized national food tables. In addition, following an 8-hour fast, participants completed a series of standardized meal challenges with controlled macronutrient composition: (a) a glucose drink (50 g of carbohydrates), (b) white bread (50 g of carbohydrates, 10 g of protein), and (c) white bread with butter (50 g of carbohydrate, 10 g of protein, 25 g of fat). In total, over 315,000 unique dish entries were recorded. Meal-response inclusion required sufficient CGM coverage to estimate both pre-meal glucose and the full 2-hour postprandial response window. Meals were excluded if additional food intake occurred within the 2-hour postprandial period (i.e. overlapping meal-responses) or if CGM data were insufficient to characterize the full glucose response window. We additionally excluded constant glucose-response arrays due to measurement errors, and meals with reported energy intake ≥2,000 kcal. We restricted the analyses to participants with >15 meals meeting all meal-level inclusion criteria. The analytic cohort included 992 participants, thus representing 50,463 free-living meals, and 4,524 standardized meals. The Food & You study received approval from the Geneva ethics commission (Ethical Approval Number: 2017-02124). The study was registered with the Swiss Federal Office of Public Health (SNCTP000002833). All participants provided informed consent before data collection. Further details of the study protocol are provided in Héritier et al. [15].

### Meal Data and Postprandial Glucose Responses

Food entries timestamped within a 30-minute period were aggregated and treated as a single food intake episode (‘*meal’*), with meal time defined as the timestamp of the first entry in the window. We adjusted meal timestamps using the closest local glucose minimum within a 30-min window preceding the recorded meal time to account for delays between food intakes and meal logging. We used the same timestamp correction in our previous PPGR analysis of the Food & You dataset [10]. For each meal, total macronutrient and micronutrient intakes, total mass, and energy intake were computed using the MyFoodRepo database. Postprandial glucose responses were characterized using four complementary outcomes derived from the 2-hour postprandial glucose response (PPGR) window. The incremental area under the curve (iAUC) was calculated relative to the pre-meal glucose baseline using the trapezoidal rule applied to 15-minute CGM measurements [16], clipping below-baseline glucose values. Maximum glucose (MaxGlu) was defined as the highest glucose level reached during the 2-hour postprandial period. Peak duration (PeakDuration) quantifies the cumulative time during which glucose levels remained above the pre-meal baseline within the same window. Finally, the 2-hour glucose level (Glu120) was defined as the glucose concentration measured at the end of the 2-hour PPGR period. Postprandial outcomes are illustrated schematically in **Figure 1a**.

**Figure 1.**
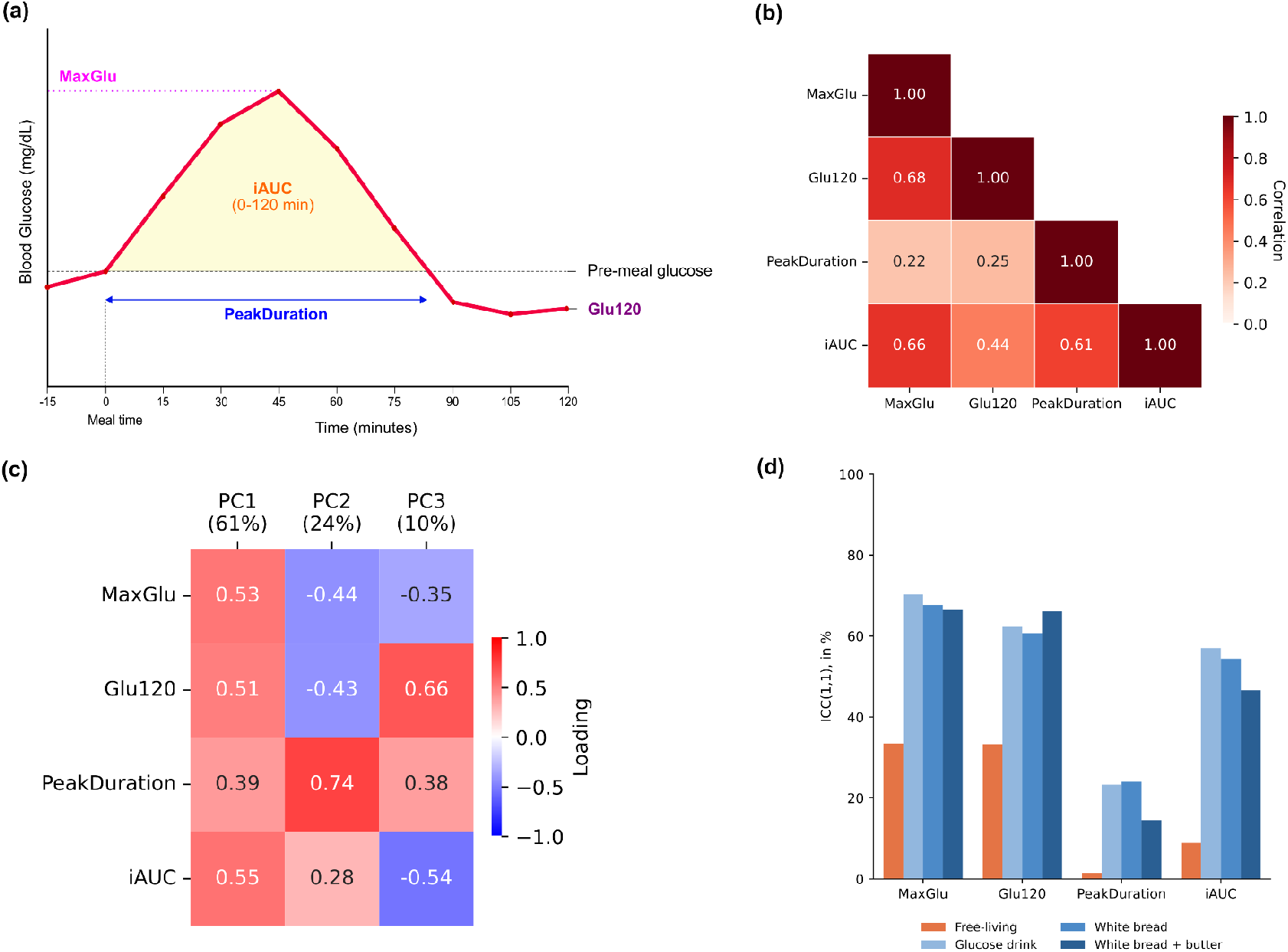
Postprandial responses are multidimensional and dominated by within-individual variability. **(a)** Schematic illustration of the four PPGR outcomes. **(b)** Heatmap of pairwise Pearson’s correlation of PPGR outcomes across all 54,987 meals, including standardized meals. **(c)** PCA loadings estimated from 50,463 free-living meal responses. **(d)** Bar plot comparing the repeatability (ICC(1,1)) of PPGR outcomes across free-living meals and standardized challenges.

Pre-meal glucose was defined as the glucose level read at the corrected meal time. Meals were included only if CGM data were available to define pre-meal glucose. Meals with CGM measurement gaps exceeding 45 minutes were excluded. Missing CGM measurements within eligible postprandial windows were completed using linear interpolation. For each meal logged at time *t*, all predictors were constructed from data available prior to logging time. Current meal features describe its nutrient content. Separate features summed nutrient intake over the previous 1, 2, 3, and 6 hours to capture recent dietary exposure. Meals were also characterized by their absolute food weights (in grams) from major food groups (*grains and pulses*; *sweets, snacks, and alcohol*; *non-alcoholic beverages*; *dairy, meat, fish, eggs, and tofu*; *vegetables and fruits*; and *oils, fats, and nuts*). We estimated the trends in past short-term glucose dynamics by calculating the least-squares slopes of CGM glucose versus time over the preceding 1, 2, 4 and 6 hours. Contextual features included the time since the last meal and the time within the tracking day. Finally, demographic and anthropometric measures (age, sex, height, weight, body mass index (BMI), waist and hip circumference) were excluded from the model training. Thus, the model input features represented information available at meal onset: immediate and recent past food intakes, the pre-meal glucose and preceding glycemic trends. The demographic and anthropometric features were excluded from the model fitting, and used post-fit.

### Multivariate mixed-effects modelling of postprandial responses

We employed a multivariate mixed-effects machine-learning framework (MMER, [17]) to separate the meal-level response surface shared across participants from participant-specific deviations. Although the MMER estimator was introduced in a different applied domain [17], this model is a general multivariate mixed-effects formulation that does not require field-specific assumptions. We used XGBoost [18] as the fixed-effects component model to estimate the non-linear meal-response surface shared across the study population. The participant-specific random effects captured the deviations from this shared surface. For each participant, we fitted a random intercept on every Z-scored PPGR outcome. These intercepts were expressed in units of the outcome SD. Random slopes were fitted for the Z-scored macronutrient predictors. Each slope measured the participant-specific deviation in each PPGR outcome, per 1-SD increase in the corresponding macronutrient. The covariance matrix τ of random effects characterised the between-participant variation across outcomes within the model. We eigendecomposed this matrix to obtain eigenvectors (‘*PC*_*Full*_’), which defined the axes of between-individual variation, and eigenvalues λ_k_ giving the variance along each axis. Effective dimensionality was quantified using the ratio d_eff_ defined as d_eff_ = (Σ_k_λ_k_)^2^/(Σ_k_λ_k_^2^) [19]. We clipped negative eigenvalues to zero because of numerical precision. We estimated the uncertainty in covariance-derived quantities using 1,000 parametric bootstrap replicates. The full mathematical and estimation details are described in the Supplementary Methods.

### Implementation

The MMER-XGBoost model was fitted with the open-source MMER Python package [20], which allows modular specification of the fixed-effects learner. To minimize scale-dependence effects, both PPGR outcomes and macronutrient predictors were Z-scored (population-level means and standard deviations) using the training set statistics before model fitting. The scaling parameters obtained during training were applied for predictions. For the linear model variants, missing predictors were imputed using participant-specific means calculated from the training data. We imputed the training-population means when a participant-specific estimate was unavailable. CGM metrics were computed using the cgmquantify package in Python [21], excluding all standardized meal response windows.

### Model comparison

We compared models that separately varied the population response surface and the degree of personalization. Linear regression provided a non-personalized linear baseline. The linear MMER (LMMER) added participant-specific random intercepts and macronutrient slopes to a linear population-level response surface. XGBoost modeled a nonlinear population response without individualization. MMER-XGBoost with random intercepts only allowed nonlinear modelling with personalization limited to overall glycemic elevation. The final MMER-XGBoost model included both random intercepts and macronutrient slopes, allowing personalization of nutrient responsiveness. The XGBoost hyperparameters inside the MMER-XGBoost framework were optimized using Optuna [22] by maximizing the R^2^ under the same cross-validation design used for the model comparison (see **Supplementary Methods**). We applied the same XGBoost hyperparameters across all XGBoost-based variants. We excluded standardized meals from the hyperparameter search and every training fold, for every participant and every model variant. The standardized meals did not contribute to the training-set statistics used for Z-scoring. Each standardized meal was assigned to the fold whose held-out block contained its timestamp and was predicted by that fold’s model. We predicted standardized meal responses from the existing fold model to evaluate the transfer to challenges excluded from the model fitting. Table 3 reports performance pooled across all 54,987 meal responses including standardized meals. We reported the prediction performance across the 4,524 standardized meals separately. Predicting them therefore requires no re-fitting: performance on standardized meals is out-of-fold prediction from models fitted without standardized meals, and evaluates transfer of individual trait estimates to a meal category absent from training.

We evaluated the predictive performance using a participant-wise, temporally blocked five-fold cross-validation. Within each participant, meals were partitioned into five contiguous, non-overlapping temporal blocks of approximately equal duration (2.8 days). One temporal block per participant was held out per fold, and models were trained on the remaining blocks. To prevent leakage due to history-based features, a 6-hour embargo was applied on both sides of each participant’s test block, excluding embargoed meals from training. This model evaluation strategy, including the temporal embargo, is described in detail and illustrated in **Supplementary Note 2**. Therefore, this cross-validation evaluates *within-individual temporal generalization* to unseen meals in previously observed individuals. In this design, training observations could precede or follow each held-out block. We also evaluated the prospective prediction performance using only earlier observations in the monitoring-length analysis. Performance was summarized by Pearson’s correlation (r), univariate and multivariate coefficients of determination (R^2^) between predicted and observed postprandial glucose response outcomes.

## Results

### Postprandial responses are multidimensional, with within-individual variability exceeding between-individual differences

The four PPGR outcomes were correlated to varying degrees (**Figure 1.b**). MaxGlu was associated with both Glu120 (Pearson’s r = 0.68) and iAUC (r = 0.66). The time spent above pre-meal glucose related only weakly to the peak height and recovery (PeakDuration vs MaxGlu: r = 0.22, Glu120: r = 0.25), and more strongly to the peak area (r = 0.61), as expected since longer excursions contribute to a larger integral. Principal component analysis of the free-living outcomes identified two leading axes that together explained 85.8% of the standardized outcome variance (**Figure 1.c**). The first axis explained 61.4% of the variance and loaded positively on all four outcomes (loadings ranging from 0.39 to 0.55), capturing overall response magnitude. The second axis explained 24.4% of the variance, and contrasted longer time above pre-meal glucose (+0.74) and greater peak areas (+0.28), with lower absolute peak and 120-minute glucose levels (−0.44 and −0.43). Positive scores on the second outcome axis indicated flatter and prolonged glucose excursions. The third axis (9.9%) contrasted Glu120 with iAUC, while the fourth axis contained only residual variation (4.3%). At the meal level, most of the variation in glucose excursions is described by their overall sizes and their recovery profiles.

We decomposed the free-living variance of each outcome into between- and within-individual components using random-intercept models, with participants as the grouping factor. We found that the within-individual component was the larger one across all outcomes (**Table 1**). Across outcomes, the within-individual SD ranged from 82% to 99% of the total marginal SD. For example, we measured a within-individual SD in iAUC of 1167 min·mg/dL against a between-individual SD of 365.5 min·mg/dL.

**Table 1.** Free-living variance decomposition by outcome. Standard deviations (SD) are reported in original outcome units. “Within-participant SD / marginal SD” represents the within-individual SD divided by the marginal population SD. The intraclass correlation coefficient ICC(1,1) represents the percentage of total variance attributable to between-participant differences. (n = 50,463 free-living meals, 992 participants).

| Outcome | Between-individual SD | Within-individual SD | Within-participant SD / marginal SD | ICC(1,1) |
| --- | --- | --- | --- | --- |
| MaxGlu (mg/dL) | 12.9 | 18.2 | 82% | 33.5% |
| Glu120 (mg/dL) | 10.5 | 14.8 | 82% | 33.3% |
| PeakDuration (min) | 4.7 | 37.7 | 99% | 1.5% |
| iAUC (min·mg/dL) | 365.5 | 1167.1 | 95% | 8.9% |

However, low free-living reproducibility does not indicate the absence of between-individual differences in postprandial response. These differences are obscured by contextual variability. Under standardized challenges (glucose drink, white bread, or white bread with butter), the intraclass correlation rose relative to free-living single meals (**Table 2**). MaxGlu reached 66.6% to 70.3% across the three challenges, Glu120 from 60.7% to 66.1%, and iAUC 46.6% to 57.0%, against free-living values of 33.5%, 33.3%, and 8.9%. The low magnitude of PeakDuration’s ICC (14.5% to 24.0%) can partly be explained by its measurement ceiling. These ICC values showed that postprandial responses varied systematically between individuals. This individuality is masked by variations in meal and glycemic context.

**Table 2.** Within-participant repeatability of glycemic outcomes. Intraclass correlation coefficients (ICC(1,1)) estimated from intercept-only random-intercept mixed-effects models with participants as the grouping factor. ICCs represent the proportion of total variance attributable to between-participant differences. Standardized meal columns report the number of participants (N) with at least one usable response for that challenge type.

| Outcome | Free-living (N=992) | Glucose drink (N=885) | White bread (N=759) | White bread + butter (N=694) |
| --- | --- | --- | --- | --- |
| MaxGlu (mg/dL) | 33.5% | 70.3% | 67.7% | 66.6% |
| Glu120 (mg/dL) | 33.3% | 62.5% | 60.7% | 66.1% |
| PeakDuration (min) | 1.5% | 23.3% | 24.0% | 14.5% |
| iAUC (min·mg/dL) | 8.9% | 57% | 54.4% | 46.6% |

**Table 3.** Model performance. Table of predictive performance across model variants and prediction targets. Multivariate R^2^ quantifies the uniform average multivariate coefficient of determination across the jointly modeled PPGR outcomes. The prediction performance of standardized meal responses was evaluated on all evaluable standardized-meal response measurements. Outcome-specific entries report R^2^ for each PPGR dimension, with Pearson’s r in parentheses. Negative R^2^ indicates prediction worse than the outcome mean.

| Models | Free-living and standardized meals<br>N= 54,987 |  |  |  |  | Standardized meals only<br>N= 4524 |  |  |  |  |
| --- | --- | --- | --- | --- | --- | --- | --- | --- | --- | --- |
| | Multivariate<br>$R^2$ | Outcome-specific $R^2$ (r) | | | | Multivariate<br>$R^2$ | Outcome-specific $R^2$ (r) | | | |
|  |  | MaxGlu | Peak<br>Duration | Glu120 | iAUC |  | MaxGlu | Peak<br>Duration | Glu120 | iAUC |
| **MMER-XGBoost**<br>Random intercept and slopes | 0.51 | 0.62<br>(0.79) | 0.44<br>(0.67) | 0.47<br>(0.69) | 0.50<br>(0.70) | 0.31 | 0.49<br>(0.72) | 0.12<br>(0.40) | 0.33<br>(0.57) | 0.31<br>(0.59) |
| **MMER-XGBoost**<br>Random intercept only | 0.50 | 0.61<br>(0.78) | 0.44<br>(0.67) | 0.46<br>(0.68) | 0.49<br>(0.70) | 0.28 | 0.44<br>(0.69) | 0.13<br>(0.40) | 0.30<br>(0.55) | 0.27<br>(0.55) |
| **XGBoost**<br>No random effects | 0.45 | 0.55<br>(0.75) | 0.43<br>(0.65) | 0.42<br>(0.65) | 0.42<br>(0.65) | 0.18 | 0.30<br>(0.59) | 0.11<br>(0.35) | 0.22<br>(0.47) | 0.09<br>(0.36) |
| **LMMER**<br>Random intercept and slopes | 0.40 | 0.50<br>(0.71) | 0.34<br>(0.58) | 0.42<br>(0.65) | 0.36<br>(0.60) | 0.00 | 0.01<br>(0.59) | -0.19<br>(0.35) | 0.32<br>(0.57) | -0.12<br>(0.45) |
| **LMMER**<br>Random intercept only | 0.39 | 0.48<br>(0.70) | 0.33<br>(0.58) | 0.41<br>(0.64) | 0.34<br>(0.59) | -0.05 | -0.05<br>(0.57) | -0.23<br>(0.35) | 0.28<br>(0.55) | -0.20<br>(0.41) |
| **Linear Regression**<br>No random effects | 0.33 | 0.41<br>(0.64) | 0.30<br>(0.55) | 0.35<br>(0.59) | 0.25<br>(0.50) | -0.19 | -0.21<br>(0.46) | -0.35<br>(0.29) | 0.20<br>(0.47) | -0.40<br>(0.11) |

### Participant-specific response patterns generalize to standardized meal challenges

Using the model family described in **Methods**, we jointly predicted the four PPGR outcomes, cross-validating within each participant with five temporal folds and no standardized meals in any training fold. All models estimated held-out response outcomes with different prediction performance (**Table 3**). MMER-XGBoost achieved the best predictive performance, with a predictive multivariate R^2^ of 0.51. In comparison, the non-individualized XGBoost achieved a predictive R^2^ of 0.45, while the linear mixed-effects model reached R^2^=0.40, and the linear regression R^2^=0.33. The predictive performance of MMER-XGBoost was strongest for MaxGlu (R^2^ = 0.62, r=0.79), while Glu120, PeakDuration and iAUC reached 0.47, 0.44 and 0.50, respectively.

We then applied the same cross-validation models to the standardized meals (glucose drinks, white-bread, and white bread with butter challenges) which were excluded from every training fold. Each challenge was predicted by the fold model whose held-out block contained it. The 6-hour embargo applied to the free-living test blocks also separates it from the training meals. In this setup, we evaluated the transfer of individual trait estimates learned from free-living meals to a meal type the models had never seen. The MMER-XGBoost model achieved the highest multivariate transfer performance, with a multivariate R^2^ of 0.31, compared with 0.28 for the random-intercept-only variant and 0.18 for non-individualized XGBoost (see: **Table 3**). The outcome-specific R^2^ values achieved by the MMER-XGBoost model were 0.49 for MaxGlu, 0.33 for Glu120, 0.31 for iAUC and 0.12 for PeakDuration.

The XGBoost model captured the nonlinear patterns and interactions missed by the linear regression model. The non-individualized XGBoost achieved a multivariate R^2^ of 0.45, compared with 0.33 for the ordinary linear regression. Adding the random effects increased the multivariate R^2^ to 0.51. The benefit of individualization was more apparent when the model predictions were evaluated on held-out standardized meals. The XGBoost explained a modest fraction of multivariate variance (multivariate R^2^ = 0.18), whereas the MMER-XGBoost reached R^2^ = 0.31. Most of the gain in predictive performance came from the random intercepts. Random slopes made a small contribution to glucose response predictions (multivariate R^2^ = 0.51 versus 0.50), but the difference in performance was more apparent in standardized meals (multivariate R^2^ = 0.31 versus 0.28).

A participant-level mixed-effects comparison confirmed that MMER-XGBoost also provided the best within-individual response prediction performance, even after adjustment for the number of glucose response records. Relative to the ordinary linear regression model, MMER-XGBoost increased participant-level multivariate R^2^ by 0.23 (95% CI: 0.21-0.24, *p* < 0.001), and exceeded the non-individualized XGBoost by approximately 0.08 R^2^ units.

We repeated the model evaluation with participants entirely held out during training (leave-participants-out, GroupKFold) as a complementary analysis. The predictive performance was lower since the model can only involve the population-level structure during the prediction. Across all meals, the linear regression achieved a multivariate R^2^ of 0.34, while its mixed-effects variant reached 0.32. In contrast, XGBoost reached a multivariate R^2^ of 0.46, while MMER-XGBoost achieved 0.43. The transfer from this cold-start setting was weaker on standardized meal predictions. The ordinary and mixed-effects linear regression models performed worse than the mean baseline prediction (R^2^ = −0.17 and −0.15, respectively). Nonlinear models performed better, but their predictive performance remained modest, with better performance from the non-individualized XGBoost (XGBoost: R^2^ = 0.21, MMER-XGBoost: R^2^ = 0.19). These results support that, by design, MMER-XGBoost predictions rely on the population-level response surface in the absence of previous observations in unseen participants.

Finally, we assessed the predictive performance of MMER-XGBoost as a function of monitoring duration. We restricted this analysis to participants with at least 14 days of available data and discarded observations beyond day 14 (N=853, 139 participants excluded). We trained the MMER-XGBoost model on the first 1 to 11 days of free-living meals and predicted a fixed held-out block comprising days 12-14. The multivariate R^2^ increased monotonically with monitoring duration. After one day, MMER-XGBoost’s performance (R^2^ = 0.290) was worse than the population-shared surface performance (R^2^ = 0.316). We observed that by day 3, the MMER-XGBoost performance was better than the population-shared response surface (0.392 vs 0.387; ΔR^2^ = 0.005). By day 11, predictions including the fitted random effects reached R^2^=0.50, compared with R^2^=0.41 for predictions from the fixed-effect component of the same MMER-XGBoost fit (ΔR^2^=0.09). The outcome-specific predictive performance followed the same patterns. The individualization improved the prediction performance after 3 days of training data for MaxGlu, Glu120 and PeakDuration, and 4 days for iAUC. Using all 11 days of data, MMER-XGBoost outperformed the population-shared predictions, with outcome-specific ΔR^2^ of approximately 0.12 for iAUC, 0.09 for MaxGlu, 0.10 for Glu120, and 0.04 for PeakDuration.

**Figure 2.**
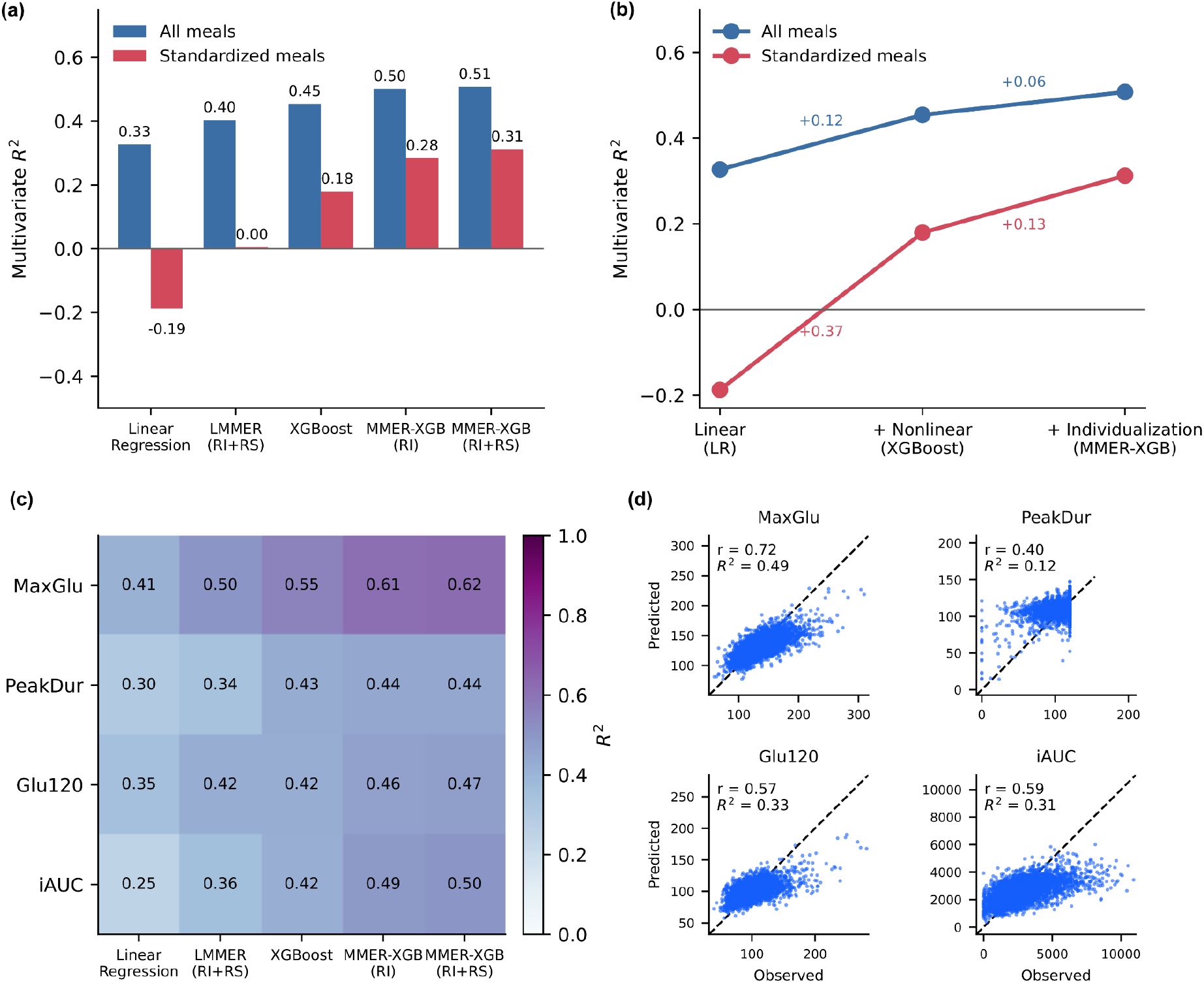
Predictive performance of postprandial glycemic response models. **(a)** Multivariate R^2^ (uniform average across the four outcomes) for each model, evaluated on all meal responses (blue) and standardized meals (red). Bars show cross-validated performance. **(b)** Decomposition of the joint predictive signal across model classes: population-level linear regression (LR), population-level nonlinear regression (XGBoost) and individualized nonlinear regression (MMER-XGB). The blue and red curves show the multivariate R^2^ across model predictions for all meals and standardized meals, respectively. **(c)** Outcome-specific R^2^ for each model on all meal responses, across maximum glucose (MaxGlu), peak duration (PeakDur), glucose at 120 min (Glu120) and positive incremental AUC (iAUC). **(d)** Scatter plots of standardized meal responses across outcomes.

### Between-individual glycemic variation is low-dimensional and reproducible, with elevated glucose responses and carbohydrate responsiveness

The principal component analysis of the random-effect covariance matrix revealed that between-individual glycemic variation was concentrated in a small number of dimensions. We denote the axes obtained from this random-effect covariance eigendecomposition as PC_Full_, to differentiate them from the other principal component analyses. PC_Full_1 explained 69.3% of the model-estimated between-participant variance (95% CI: 64.1-71.0%), and PC_Full_2 explained 11.8% (95% CI: 10.2-13.5%). These two axes accounted for 81.1% (95% CI: 76.2-82.4%) of the total variance. A third component explained 5.0% (95% CI: 4.5-6.3%), increasing the cumulative variance explained by these first three axes to 86.2% (95% CI: 82.1-87.4%). The effective dimensionality was 2.00 (95% CI: 1.92-2.30). Since the random-effect space comprises 20 coordinates (five terms across four outcomes), an effective dimensionality of 2 represents a large compression of the model’s parametrization. Bootstrap alignment confirmed that the first two eigenvectors were highly persistent (median loading cosine similarity: PC_Full_1 = 0.999, PC_Full_2= 0.992), whereas the third was moderately stable (PC_Full_3 = 0.909). This dominant covariance structure was also robust to the definition of the PPGR outcomes. When MaxGlu and Glu120 were instead expressed as changes from pre-meal glucose (ΔMaxGlu, and ΔGlu120), PC_Full_1 and PC_Full_2 explained 67.1% and 11.0% of between-participant variance, respectively, compared with 69.3% and 11.8% in the original specification. The corresponding loading vectors were nearly identical (cosine similarity = 0.996 and 0.992), and participant scores remained highly concordant (Pearson’s r = 0.998 for PC_Full_1 and 0.975 for PC_Full_2; Supplementary Note 8).

PC_Full_1 had elevated positive loadings on all random intercepts. Modest carbohydrate-responsiveness loadings were aligned with this axis (see: ***Table 4***). We interpreted that individuals scoring highly on PC_Full_1 had generally large postprandial excursions. The PC_Full_2 axis (11.8% of between-individual variance) combined carbohydrate responsiveness with lower late glucose elevations. Carbohydrate slopes loaded positively on every outcome (e.g. iAUC: +0.51), while random intercepts in Glu120 and PeakDuration loaded negatively (respectively −0.48, and −0.32).

**Table 4.** Principal component loadings of participant-specific glycemic random effects. Loading description of the two leading principal components of the participant-level random-effect covariance matrix. For each component, we reported the ten largest loadings ranked by absolute value. Values in brackets indicate 95% bootstrap confidence intervals.

| PC<sub>Full</sub>1 (69.3% of variance explained) |  |  | PC<sub>Full</sub>2 (11.8% of variance explained) |  |  |
| --- | --- | --- | --- | --- | --- |
| Random effect | PPGR outcome | Loadings | Random effect | PPGR outcome | Loadings |
| Intercept | iAUC | +0.54 [+0.53; +0.55] | Carbohydrates | iAUC | +0.51 [+0.48; +0.53] |
| Intercept | MaxGlu | +0.46 [+0.45; +0.47] | Intercept | Glu120 | -0.48 [-0.54; -0.40] |
| Intercept | Glu120 | +0.44 [+0.42; +0.46] | Carbohydrates | MaxGlu | +0.39 [+0.35; +0.41] |
| Carbohydrates | iAUC | +0.33 [+0.31; +0.35] | Carbohydrates | Glu120 | +0.35 [+0.28; +0.41] |
| Intercept | PeakDuration | +0.28 [+0.27; +0.30] | Intercept | PeakDuration | -0.32 [-0.34; -0.26] |
| Carbohydrates | MaxGlu | +0.25 [+0.24; +0.27] | Fiber | iAUC | -0.18 [-0.24; -0.11] |
| Carbohydrates | Glu120 | +0.20 [+0.19; +0.23] | Carbohydrates | PeakDuration | +0.18 [+0.15; +0.23] |
| Protein | iAUC | -0.03 [-0.05; -0.01] | Fiber | MaxGlu | -0.14 [-0.20; -0.09] |
| Protein | MaxGlu | -0.03 [-0.04; -0.01] | Fat | iAUC | -0.11 [-0.18; -0.05] |
| Fiber | iAUC | -0.02 [-0.05; -0.01] | Fiber | Glu120 | -0.10 [-0.16; -0.05] |

Then, we calculated the Schur complement of *τ* against the four-intercepts covariance to isolate the covariance of macronutrient responsiveness (see: **Supplementary Note 3**). The effective dimensionality of the Schur complement was 4.31 (95% CI: 4.07-5.56). The leading axis PC_C_1 mainly described carbohydrate-versus-fiber responsiveness, explaining 37.2% of residual variance (95% CI: 29.1-42.1%), with positive carbohydrate loadings on iAUC (+0.54), MaxGlu (+0.48), and Glu120 (+0.40), and negative fiber loadings on iAUC (−0.34), MaxGlu (−0.28), and Glu120 (−0.22). PC_C_1 was highly stable through bootstrap iterations (median cosine similarity: 0.972). This observation supports that carbohydrate responsiveness remains identifiable, even after accounting for an individual’s overall glycemic elevations. The second component PC_C_2 explained 24.0% of the conditional covariance (95% CI: 18.0-26.1%, median cosine similarity: 0.946) and described a protein-versus-fat contrast. These two axes captured 61.2% (95% CI: 52.0-63.1%) of the Schur complement’s variance. The third axis separated fiber-from fat-associated responsiveness, and explained 17.2% of residual variance (95% CI: 12.0-18.3%, median cosine similarity: 0.955). The fourth axis explained 5.0% of residual variance (95% CI: 4.9-7.4%) and showed weaker bootstrap alignment (median cosine similarity: 0.821).

Next, we performed two split-half reliability analyses to evaluate the estimation stability of individual characteristics. In the first split-half setting, odd- and even-indexed free-living meals formed non-overlapping subsets spanning the same monitoring period; the subsets could still share temporal, behavioral and sensor-related variation. In the second split-half setting, we chronologically divided meals into the first and second halves of each participant’s monitoring period, providing a more stringent test based on temporally separated observations. Then, we fitted the MMER-XGBoost model on each subset. We analysed the resulting participant-specific random effects by projecting them onto the principal component basis obtained from the full-data covariance point estimate to ensure a fair comparison of each estimation. As illustrated in **Figure 3d**, the dominant axis PC_Full_ 1 showed strong split-half reliability across both validation strategies (odd/even splits: Pearson’s r = 0.80, ICC(2,1) = 0.80; temporal split: r = 0.74, ICC(2,1) = 0.73). The second axis was less reliable and degraded further under temporal separation (odd/even: r= 0.47, ICC(2,1) = 0.47; temporal split: r= 0.36, ICC(2,1) = 0.35). The results obtained from these split-half analyses support the persistence of the PC_Full_1 scores over the observed monitoring period. PC_Full_2 analyses provided lower agreement and indicated lower estimation precision. As an additional check, we repeated the random-effect covariance eigendecomposition separately within each split-half setting. The low-dimensional structure was preserved across splitting schemes, with effective dimensionalities of 2.4 and 2.3 (odd/even) and 2.05 and 2.33 (temporal). As in the full data fit, PC_Full_ 1 explained 62.6 to 68.3% of variance in each split-half setting, while PC_Full_ 1 and PC_Full_ 2 explained together 73.5 to 80.5%. In addition, the eigenvector alignment was high for both axes, with PC_Full_ 1/PC_Full_ 2 cosine similarities of respectively 0.99/0.97 for odd-even splits and 0.95/0.91 for temporal splits. These results indicate that the estimated low dimensionality is itself reproducible across non-overlapping meal subsets, while reliable participant-level estimation is currently supported only for the PC_Full_1 axis.

**Figure 3.**
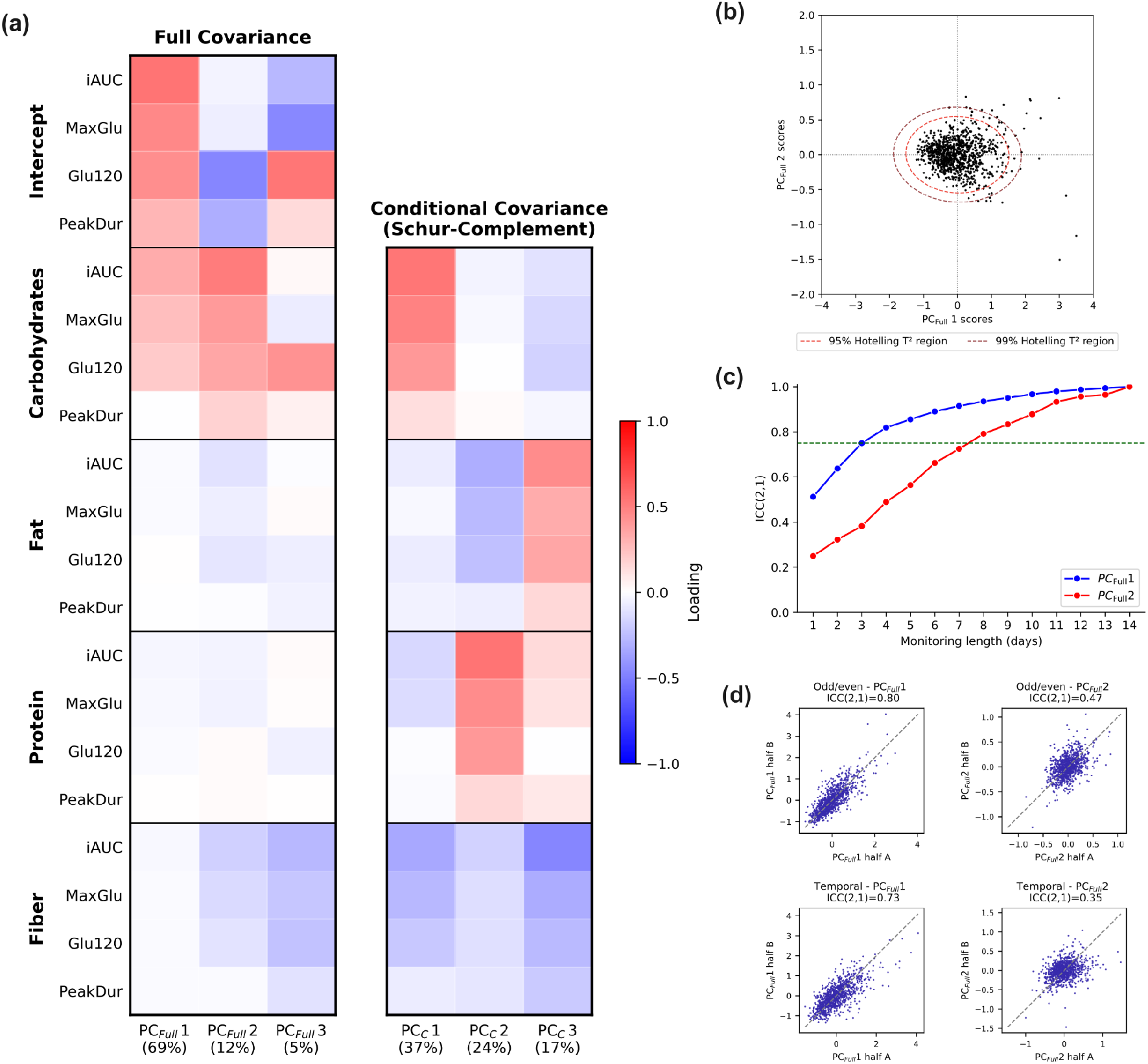
Principal axes of between-individual glycemic variation. **(a)** Heatmap of PC_Full_ loadings from the MMER-XGBoost random-effects covariance and the Schur-complement covariance (PC_C_). The variance explained by each axis is reported as percentages. **(b)** Scatter plot of participant positions in the PC_Full_ space. The inner and outer ellipses show the 95% and 99% Hotelling’s T^2^ regions, respectively. **(c)** Absolute agreement, ICC(2,1), between scores estimated from the first d monitoring days and an overlapping 14-day reference in 853 participants. The green horizontal line marks ICC = 0.75. Agreement equals 1 at day 14 by construction. **(d)** Split-half agreement in participant scores for PC_Full_ 1 and PC_Full_ 2 between the two halves of each split, evaluated using odd-even and temporal splitting schemes.

Among 853 participants with at least 14 observed monitoring days, we estimated PC_Full_1 and PC_Full_2 scores from the first 1 to 14 days and compared them with estimates from the same participants’ 14 days. Absolute agreement for PC_Full_1 increased from ICC(2,1) = 0.512 after one day to approximately 0.75 after three days and 0.82 after four days. PC_Full_2 exceeded ICC thresholds of 0.75 and 0.90 on day 8 (ICC(2,1) = 0.79) and day 11 (ICC = 0.93), respectively. A sensitivity analysis using an uncapped full-record reference fitted in all 992 participants yielded the same threshold-crossing days, with absolute-agreement ICCs differing by at most 0.038 between references (**Supplementary Note 4**). These results support early approximation of PC_Full_1 from three monitoring days, with slower convergence of PC_Full_2 toward the overlapping reference estimates.

Finally, we evaluated the transferability of a phenotype basis learned in the training participants and applied to held-out participants, without re-fitting the model. Participants were assigned to five mutually exclusive folds; in each iteration, the model was fitted using four folds and scores were inferred for participants in the remaining fold. Within each held-out fold, meal-responses were divided into odd- and even-indexed subsets, from which we separately estimated their PC_Full_ scores using the frozen phenotype basis learned from the training population. We observed that the PC_Full_1 scores showed high split-half reliability, whereas PC_Full_2 estimation reliability was more modest. Alternating meal subsets gave pooled ICC(3,1) values of 0.81 (95% CI: 0.79-0.83) for PC_Full_1 and 0.52 (95% CI: 0.48-0.57) for PC_Full_2. Chronologically separated subsets returned corresponding values of 0.74 (95% CI: 0.71-0.77) and 0.48 (95% CI: 0.44-0.53). The full methods and results are reported in **Supplementary Note 7**.

### Conventional participant characteristics explain little of the recovered response axes

Among 989 participants with complete covariates, we investigated associations of demographic and anthropometric variables with the PC_Full_ coordinates. Age and sex (sex coded ‘male’ = 1) together explained 10.2% of the variance in PC_Full_1. Adding BMI, waist, and hip circumferences increased the variance explained to 12.0% (ΔR^2^ = 0.019, nested F-test p = 1.3 ×10^−4^), showing that anthropometric measures add a small and significant amount of additional information beyond age and sex only. Age was the characteristic most strongly correlated with PC_Full_1 with a standardized β of 0.33 (p<0.001). Males scored 0.23 SD higher on PC_Full_1 than females (standardized β = 0.23, p<0.01). Adiposity metrics did not reach significance in the joint model (p>0.07). Overall, demographic and anthropometric variables explained only a small fraction of PC_Full_1 variance.

Demographic and anthropometric characteristics explained little variation in PC_Full_2. The full model accounted for 1.3% of its variance (p=0.023), with a weak positive association with age (standardized β=0.082, 95% CI: 0.013-0.151, p=0.020). Adding BMI, waist and hip circumferences to age and sex provided negligible additional explanatory power (ΔR^2^=0.0008, nested F-test p=0.85).

We evaluated PC_Full_ score predictions using five repeats of shuffled 10-fold cross-validation. The baseline linear model included age, sex and BMI. The expanded model additionally included height, weight, waist and hip circumference, waist-to-hip ratio and waist-to-height ratio. The predictive performance remained weak for both scores and models, and adding adiposity features increased the out-of-sample R^2^ from 0.10 to 0.11 for PC_Full_1. The predictions in PC_Full_2 were worse than predicting the mean for both blocks (R^2^<0.0, **Figure 4.a**). Next, we fitted a gradient-boosted model (HistGradientBoostingRegressor, [23]) to capture the potential nonlinear associations and interactions between participant characteristics and PC_Full_ scores. The gradient-boosted model achieved a mean cross-validated R^2^ of 0.072 for PC_Full_1, below the expanded linear model. Its performance was however significantly above the permutation null (null mean R^2^ = −0.11, p = 0.001). The same features explained essentially no variance in PC_Full_2, with an out-of-sample R^2^ of −0.12, and did not exceed the permutation null (p = 0.64). These results indicate that participant characteristics cannot accurately predict PC_Full_ scores.

**Figure 4.**
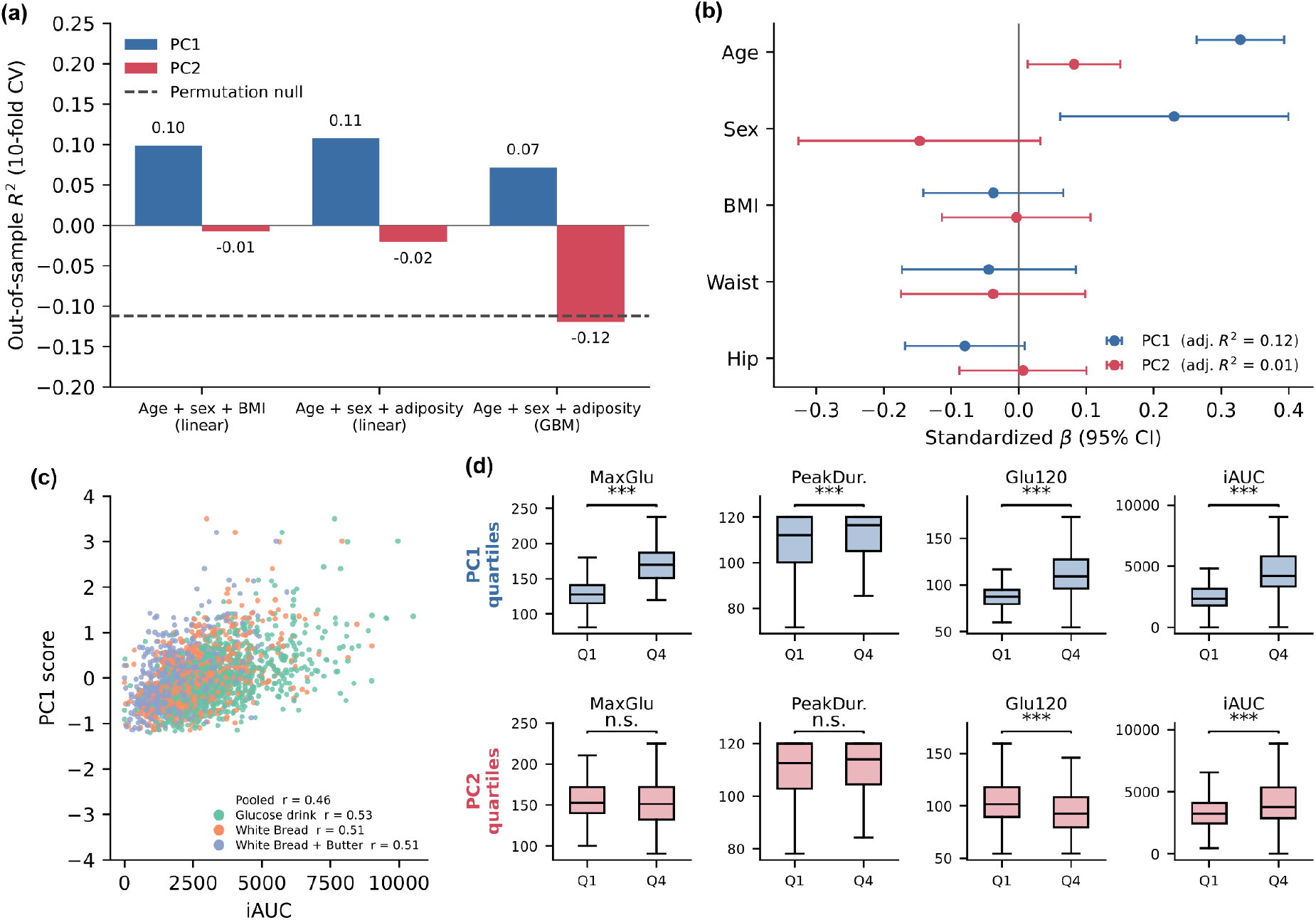
Associations of PC_Full_ scores with demographics, anthropometry, and standardized meal challenges. **(a)** Out-of-sample predictive performance of PC_Full_ scores by feature set. Significance for the gradient-boosted models was assessed using 1,000 outcome permutations and the same repeated-cross-validation statistic for the observed and permuted outcomes. **(b)** Standardized multivariable regression coefficients (β, 95% CI) for age, sex, BMI, waist, and hip. **(c)** Free-living PC_Full_1 vs standardized-challenge iAUC, pooled across challenges (coloured by type). Pooled and challenge-specific Pearson’s r associations. **(d)** Quartiles of each axis compared on four outcomes from the glucose-drink challenge (50 g of glucose); Q1 and Q4 were compared using two-sided Mann-Whitney U tests, with Benjamini-Hochberg correction across the 24 challenge-by-coordinate-by-outcome comparisons (Supplementary Table S6.3; *pFDR<0.05, **pFDR<0.01, ***pFDR<0.001; ns, not significant).

### The dominant axis generalizes to standardized glucose challenges

Both phenotype coordinates were associated with conventional CGM metrics after adjustment for age, sex, and BMI, and FDR corrections. PC_Full_1 was positively associated with overall glycemic levels and variability. The strongest associations were with average intraday SD (r_partial_ = 0.67, 95% CI: 0.63-0.70), the maximum glucose reached during the monitoring period (r_partial_=0.68, 95% CI: 0.64-0.71), and mean glucose (r_partial_=0.63, 95% CI: 0.59-0.67). We also found strong associations between PC_Full_1 scores and glucose levels at food intakes (mean pre-meal glucose levels r_partial_=0.63, 95% CI: 0.59-0.67). In contrast, PC_Full_2 showed the opposite patterns for level metrics. PC_Full_2 was associated with lower glucose levels (Q1 glucose : r_partial_= −0.61, 95% CI: −0.65- −0.57, Q3: r_partial_ = −0.50, 95% CI: −0.54 - −0.45). Participants with high PC_Full_2 scores had lower glucose levels but experienced peaks comparable to the rest of the population, resulting in large responses relative to their usual glucose levels.

We next studied the associations between PC_Full_ coordinates and standardized meal responses beyond demographics and glucose profile. In this analysis, we excluded 107 participants without valid glucose drink responses and retained 885 participants. In nested linear regression models adjusted for age, sex, BMI, mean glucose, and coefficient of variation of glucose levels, adding PC_Full_1 improved model fit for all four outcomes (all p_FDR_ <10^−10^). PC_Full_1 substantially increased the adjusted R^2^ by 0.14 for MaxGlu, and 0.17 for iAUC. The additional explanation in glucose drink variance was modest for both Glu120 (Δadjusted R^2^ =0.04) and PeakDuration (Δadjusted R^2^ =0.05). PC_Full_2 added slightly less information, increasing the adjusted R^2^ by 0.02 for both MaxGlu and iAUC (both p_FDR_<10^−5^). The PC_Full_2 did not significantly increase the variance explained in Glu120 (Δadjusted R^2^= 0.00, p_FDR_=0.75) or PeakDuration (Δadjusted R^2^=0.00, p_FDR_=0.16). Thus, PC_Full_1, and more modestly PC_Full_2, explained variance of the glucose-drink response that was not described by demographic characteristics, mean glucose or glucose variability. Corresponding analyses of the white-bread and white-bread-plus-butter challenges are reported in **Supplementary Note 6**.

Associations between PC_Full_ scores and glucose-drink responses were also evident when comparing the extreme quartiles: Q1 (N = 222), representing the lowest scores, and Q4 (N= 221), representing the highest scores (**Figure 4.d**). Participants in the highest versus lowest PC_Full_1 quartiles had higher median MaxGlu (Q4: 169.6 versus Q1: 127.6 mg/dL, p_FDR_<0.001), Glu120 (Q4: 109.3 versus Q1: 87.1 mg/dL, p_FDR_<0.001) and iAUC (Q4: 4,209.9 versus Q1: 2,333.8 min·mg/dL, p_FDR_<0.001), with a modestly longer PeakDuration (Q4: 116.4 versus Q1: 112.1 min, p_FDR_<0.001). For PC_Full_2, the highest quartile had lower median glucose at 120 minutes (Q4: Glu120: 92.3 versus Q1: 101.6 mg/dL, p_FDR_<0.001) but larger peak areas (Q4: iAUC: 3,761.4 versus Q1: 3,212.8 min·mg/dL, p_FDR_<0.001). MaxGlu and PeakDuration did not differ between quartiles (p_FDR_>0.1).

## Discussion

A central challenge in precision nutrition is that postprandial glucose responses in free-living conditions are simultaneously individualized and highly variable. The within-individual variability in PPGR outcomes exceeded the variation between individuals. Under standardized settings, we observed a higher repeatability in glucose responses, especially in glucose peaks following glucose drinks (ICC(1,1) = 0.70). In contrast, raw everyday glucose responses provide an imprecise estimate of an individual’s usual glucose response (e.g. iAUC: ICC(1,1)<0.10). These results are consistent with previous observations that glucose responses from controlled meal intakes vary in adults without diabetes [24].

Once meal and glycemic contexts are modelled through a population-shared response surface, the participant-specific deviations occupy a narrow space. In this study, we observed that 81.1% of the model-estimated variation between participants was concentrated in two axes, with one dominant axis representing 69.3% of the total variance. This axis reads most simply as overall glycemic elevation. Higher scores along this axis reflected larger glucose excursions, after adjustment for the measured meal and glycemic contexts. A recent study analyzing glucose responses from dinners found a comparable structure, with a leading axis reflecting general elevations in glucose responses [13]. The PC_Full_1 scores carried information that the conventional characteristics did not. In models already containing age, sex, BMI, mean and coefficient of variation of glucose levels, adding PC_Full_1 scores increased the adjusted R^2^ by 0.14 for glucose-drink MaxGlu, and by 0.17 for iAUC. We observed comparable gains with glucose responses from white bread and white bread with butter. This dominant axis was preserved even after expressing both MaxGlu and Glu120 relative to the pre-meal glucose. The positive association between PC_Full_1 and age is consistent with the well-described decline in postprandial glucose control with age [25], [26]. We also observed that male participants had higher PC_Full_1 scores, which suggests a sex-related difference in glucose regulation. However, demographics and anthropometrics provided limited information about an individual’s PC_Full_1 scores by explaining only 12% of PC_Full_1 score variance.

The second axis PC_Full_2 combines carbohydrate responsiveness with lower late glucose elevations and shorter glucose response duration. This axis explained a small portion of between-individual deviations (11.8%). Random slopes contributed modestly to predictive performance, and individual PC_Full_2 scores were less reproducible than PC_Full_1 scores. The lower glucose levels at 120 minutes observed in participants with higher PC_Full_2 scores should therefore not be interpreted as evidence of faster glucose clearance or favorable metabolic regulation. Adiposity features and standardized challenges were weakly associated with this axis.

We found strong reproducibility in PC_Full_1 estimations under separation of the monitoring period, whereas PC_Full_2 remained less reliable. This application requires observations from the individual being characterized. In the absence of such observations, non-individualized XGBoost performed better than MMER, showing that the benefit of the framework lies in estimating individual response patterns from repeated meals. Participant cross-fitting supported the recovery of PC_Full_1 from models fitted in other participants from the same cohort. We split the held-out participant meals into alternating subsets and estimated their PC_Full_ scores twice, and observed ICCs of 0.81 for PC_Full_1 and 0.52 for PC_Full_2. When we chronologically split meals into subsets, we measured ICCs of 0.74 and 0.48, respectively.

We observed that three monitoring days provided an early approximation of the PC_Full_1 scores obtained from the overlapping 14-day record. Three days of monitoring was also the shortest duration at which individualized prediction performance exceeded the population-shared response surface, although the gain at this point was small (ΔR^2^=0.005). These results identify approximately three days as an early transition point at which PC_Full_1 becomes concordant with longer-record estimates (ICC(2,1) ≥0.75) and individualization first begins to improve prospective prediction. PC_Full_2 scores converged more slowly and approached comparable agreement after one week of monitoring.

The cohort included adults in Switzerland without diagnosed diabetes and we could not determine whether the same variance geometry holds under impaired regulation. Dysglycemia could preserve it or reorganize it as regulatory mechanisms decouple. Without further clinical measurements, we could not identify what physiologically drives the glucose response phenotypes. Differences in insulin sensitivity and beta-cell compensation are possible contributors [11], and resolving this will require insulin measurements. External validations with individuals with diagnosed impaired glucose control, and insulin measurements are required. The CGM sensors measured interstitial glucose at 15-minute resolution, with known errors [27]. Dietary inputs came from participant logging and human annotation, so errors in macronutrient reports may have propagated into the slope terms. The PC_Full_2 had lower variance and is the axis most exposed to that noise. The glycemic phenotype space is conditional on our parametrization of one intercept and four macronutrient slopes per outcome. A richer slope structure or PPGR outcomes could resolve further axes. Within these limits, routine food intake and CGM records from everyday technologies support measurement of a reproducible dominant glycemic response coordinate and provide a candidate measure for research stratification.

## Data availability

The analysis code and results are available on GitHub at https://github.com/digitalepidemiologylab/postprandial-glucose-traits-analyses, and archived on Zenodo (DOI:10.5281/zenodo.22677786). The raw Food & You data will soon be publicly released. The link will be posted at digitalepidemiologylab.org.

## Ethics statement

We used data from the Food & You study, which received ethical approval from the Geneva ethics commission that reviewed and authorized the project (Ethical approval number: 2017-02124). The study is registered on the website of the Swiss Federal Office of Public Health (SNCTP000002833) and the platform clinicaltrials.gov (NCT03848299). All participants provided informed consent before data collection. Study procedures followed applicable local legislation and institutional requirements.

