## Supplementary Material for "Short-term postprandial glucose monitoring reveals stable traits from noisy free-living meals"

**M. Toumi[1], M. Salathé[1]**
[1]*Digital Epidemiology Lab, School of Life Sciences, School of Computer and Communication Sciences, EPFL, Lausanne, Switzerland*

### Supplementary

**Table of contents** **1**

Supplementary Methods | MMER-XGBoost model formulation, estimation, tuning, and uncertainty 1
Supplementary Note 1 | Cohort description 5
Supplementary Note 2 | Temporally blocked within-participant cross-validation and leakage control 9
Supplementary Note 3 | Macronutrient responsiveness after accounting for overall glycemic elevation 10
Supplementary Note 4 | Recoverability of participant coordinates as a function of monitoring length 12
Supplementary Note 5 | Associations between glycemic phenotype coordinates and CGM descriptors 14
Supplementary Note 6 | Concordance of glycemic phenotype coordinates and standardized meal challenge responses 16
Supplementary Note 7 | Cross-fitted reproducibility of glycemic phenotype coordinates in held-out participants 20
Supplementary Note 8 | Robustness of the glycemic phenotype to baseline-relative outcome definitions 22
References 23

#### Supplementary Methods | MMER-XGBoost model formulation, estimation, tuning, and uncertainty

**Model specification**: We modeled the four postprandial outcomes jointly within a multivariate mixed-effects regression (MMER) framework [1], using XGBoost [2] as the nonparametric fixed-effects learner.
For a participant i, $i \in [\![1, N]\!]$, and participant-specific meal j, $j \in [\![1, n_i]\!]$, where $n = \sum_i^N n_i$ represents the total number of free-living meals , and the vector of respective PPGR outcomes $Y_{ij} = [\, y_{MaxGlu,\,ij}\,, y_{Glu120,\,ij}, y_{PeakDuration,\,ij}, y_{iAUC,\,ij}\,]$, the model is defined as :

$$Y_{ij} = f_{XGB}(X_{ij}) + B_i z_{ij} + \varepsilon_{ij}$$

where $\mathbf{f_{XGB}}$ represents the population response to food and glycemic contexts, and $\mathbf{z_{ij}}$ contains for each outcome an intercept and standardized carbohydrates, fat, protein, and fiber intakes. The 4×5 matrix $\mathbf{B_i}$ contains participant-specific deviations, i.e. one intercept and four macronutrient slopes for each outcome.

The 20 participant-specific coefficients followed a zero-mean multivariate Gaussian distribution with covariance $\tau$. Meal-level residuals followed a zero-mean multivariate Gaussian distribution with covariance $\phi$. Random effects were independent across participants and independent of residuals, which were assumed independent across meals. The covariance matrices captured dependence within and across outcomes, and were estimated during the MMER-XGBoost fitting procedure.

The models were fitted using the MMER package's EM-based procedure [1], alternating between estimating participant random effects and refitting XGBoost to responses adjusted for these effects, and updating the covariance matrices. We fitted XGBoost using outcome-specific trees (strategy = "*one_output_per_tree*"). The entire MMER-XGBoost model was fitted until convergence, with a relative likelihood tolerance of $10^{-7}$ or 50 iterations as stopping criteria.

**Hyperparameter tuning**. The XGBoost hyperparameters were optimized using 200 trials of sequential model-based optimization with Optuna's Tree-structured Parzen Estimator (TPE, random seed= 42). Each candidate configuration was evaluated by refitting the complete MMER framework under the five-fold temporally blocked CV. We selected hyperparameters by maximizing the multivariate $R^2$ across the four jointly modeled PPGR outcomes. The selected hyperparameters were subsequently fixed across all XGBoost-based variants in the model comparison.

| Hyperparameters | Search space | Scale | Selected values |
| --- | --- | --- | --- |
| n_estimators | [600, 1200] (step 100) | integer | 1200 |
| learning rate | [0.01, 0.5] | log-uniform | 0.02258140698292982 |
| max_depth | [4, 10] | integer | 6 |
| subsample | [0.7, 1.0] | uniform | 0.7743798042397052 |
| colsample_bytree | [0.7, 1.0] | uniform | 0.7166838990816966 |
| reg_alpha | [$10^{-3}$, 10] | log-uniform | 2.3759453153584325 |
| reg_lambda | [$10^{-3}$, 10] | log-uniform | 0.031006288561511164 |
| min_child_weight | [1, 20] | integer | 8 |
| gamma | [0, 5] | uniform | 0.6335343145218796 |

***Supplementary Methods Table 1. Search space and selected hyperparameters for the XGBoost learner of MMER.*** *Hyperparameter search space and best hyperparameter set found after 200 trials of Optuna's Tree-structured Parzen Estimator (random seed = 42). Each trial refitted the complete MMER pipeline under the five-fold temporally blocked cross-validation with a 6-h embargo (presented above). The objective was maximizing the uniform-average $R^2$ across the four jointly modeled PPGR outcomes (positive iAUC, peak glucose, peak duration, end glucose), calculated on the pooled out-of-fold predictions. The selected configuration reached a multivariate $R^2$ of 0.51. Fixed settings not tuned: booster = 'gbtree', tree_method = 'hist', multi_strategy = 'one_output_per_tree', seed = 42. All other parameters were set at library defaults.*

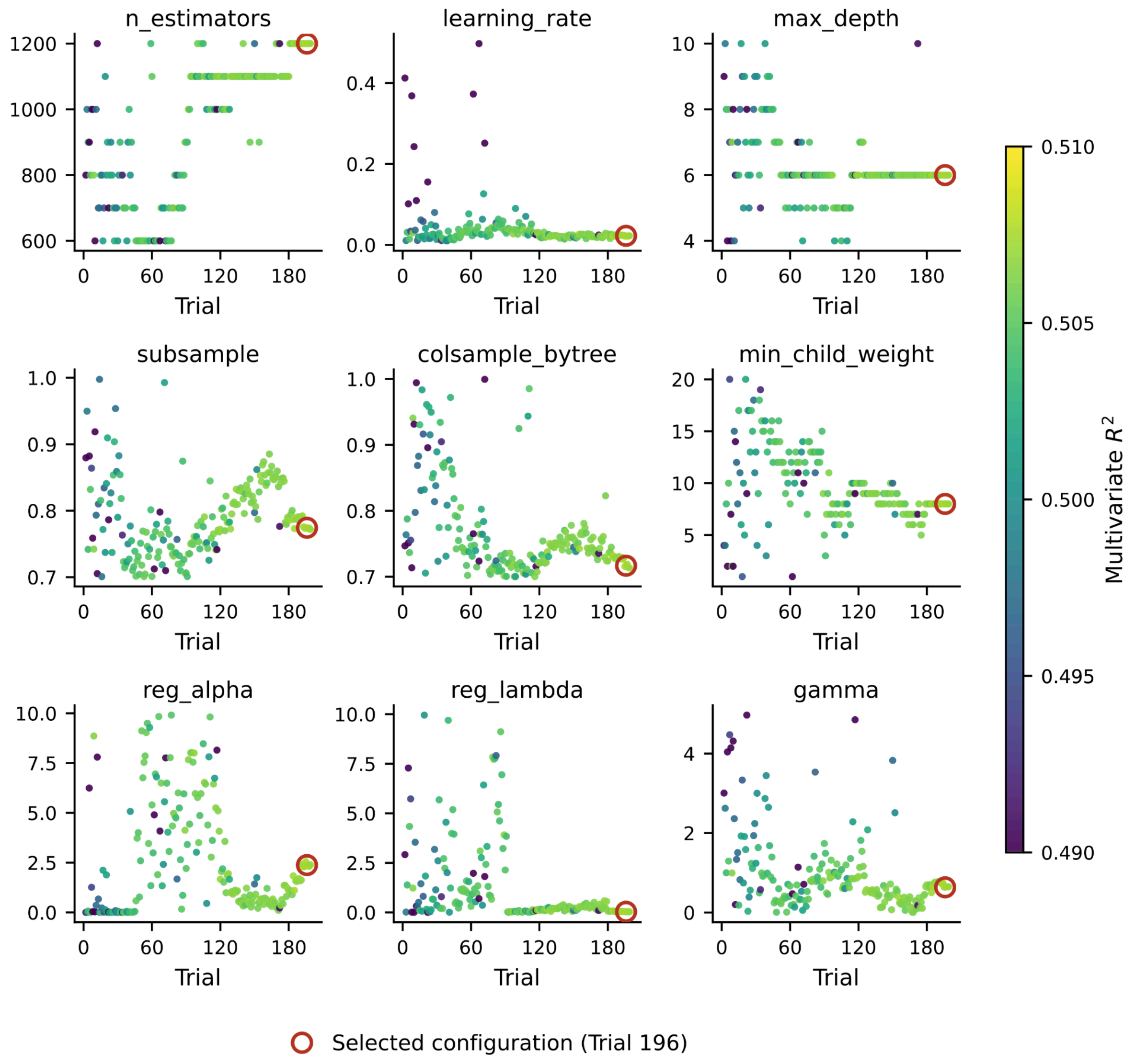


***Supplementary Methods Figure 1. Hyperparameter values sampled across trials****. Values sampled by the Tree-structured Parzen Estimator at each trial, coloured by the corresponding multivariate $R^2$. Red rings mark the selected configuration (see:* ***Supplementary Methods Table 1****)*

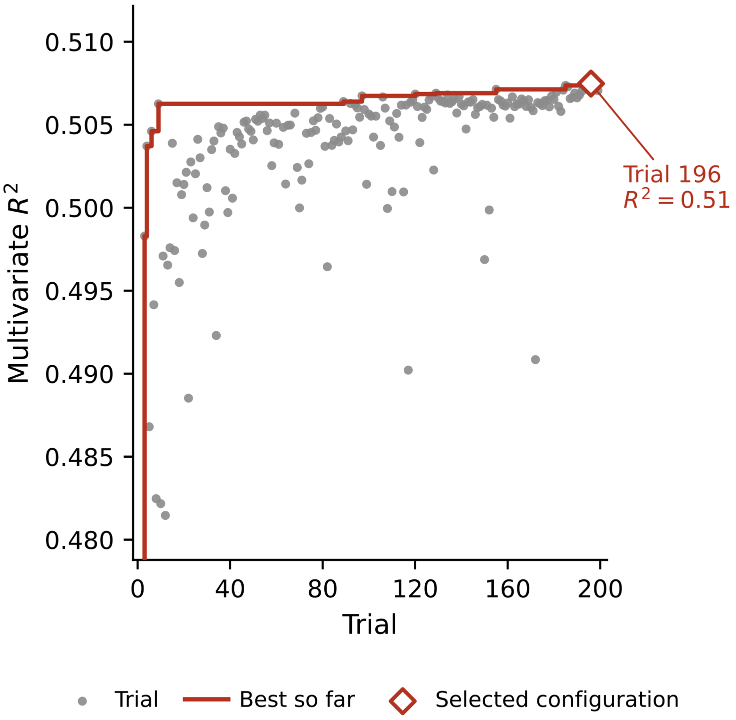


***Supplementary Methods Figure 2. XGBoost hyperparameter search.*** *Objective value per trial (grey) and running best (red) over the 200 Optuna trials. The objective is the multivariate (uniform-average) $R^2$ across the four jointly modelled PPGR outcomes under five-fold temporally blocked cross-validation with a 6-h embargo, with the complete MMER framework refitted for each trial. The selected configuration (open diamond) reached $R^2 = 0.51$.*

**Uncertainty estimation.** We quantified the uncertainty in the random-effect covariance structure using 1,000 parametric bootstrap replicates. For each replicate, we retained the observed meal predictors, participant grouping, and scaling parameters. Then, we simulated new outcomes from the fitted model using Gaussian participant random effects and meal-level residuals drawn from their estimated distributions. We refitted the entire MMER-XGBoost model, including the XGBoost learner, and repeated the eigendecomposition, to extract the principal component loadings, explained variance, and effective dimensionality. Bootstrap components were matched to the original components by direction and sign before summarizing their loadings. We calculated 95% confidence intervals from the 2.5th and 97.5th percentiles of bootstrap estimates.

#### Supplementary Note 1 | Cohort description

The study population comprised 992 individuals without diagnosed diabetes. Baseline participant characteristics are summarized in **Supplementary Table S1.1**.

| **Characteristics** | **All participants (N=992)** |
|---|---|
| Age, years | 39.7 ± 12.1 |
| sex, n (%) | Male: n=429 (43%), Female: n=563 (57%) |
| BMI, kg/m$^2$ | 23.6 ± 3.6 |
| Cohort, n (%) | Cohort B: 848 (85%), Cohort C: 144 (15%) |
| Observation length, days | 15.6 ± 5.1 |
| Recorded meals per participant, n | All meals: 55.4 ± 18.9<br>Ad libitum meals: 50.9 ± 17.7<br>Standardized meals: 4.7 ± 2.2 |

***Supplementary Table S1.1: Baseline characteristics of the study population**. Values are reported as mean ± standard deviation (SD) unless otherwise specified. Standardized-meal counts are summarized among the 963 participants with at least one standardized meal; other participant summaries use the denominators indicated in the table. Observation length counts dates with evaluable records.*

| **Category** | **Feature** | **Unit** | **Mean ± SD** |
|---|---|---|---|
| Meal composition | Carbohydrate | g | 34.48 ± 35.16 |
| Meal composition | Fat | g | 14.46 ± 16.85 |
| Meal composition | Protein | g | 12.21 ± 16.16 |
| Meal composition | Dietary fiber | g | 3.79 ± 4.79 |
| Meal composition | Meal energy | kcal | 331.02 ± 321.26 |
| Meal composition | Meal mass | g | 284.95 ± 233.06 |
| Meal composition | Estimated net carbohydrate | g | 30.72 ± 32.23 |
| Meal composition | Estimated sugar fraction | Ratio | 0.47 ± 1.14 |
| Meal composition | Alcohol | g | 0.94 ± 4.28 |
| Meal composition | Beta-carotene | g | 4.13e-4 ± 1.43e-3 |
| Meal composition | Calcium | g | 9.46e-2 ± 1.63e-1 |
| Meal composition | Cholesterol | g | 4.03e-2 ± 9.89e-2 |
| Meal composition | Monounsaturated fatty acids | g | 3.42 ± 29.64 |
| Meal composition | Polyunsaturated fatty acids | g | 1.47 ± 7.00 |
| Meal composition | Saturated fatty acids | g | 5.38 ± 6.96 |
| Meal composition | Folate | g | 2.75e-5 ± 4.53e-5 |
| Meal composition | Iron | g | 1.20e-3 ± 3.23e-3 |
| Meal composition | Magnesium | g | 3.16e-2 ± 4.40e-2 |
| Meal composition | Niacin | g | 1.55e-3 ± 3.10e-3 |
| Meal composition | Pantothenic acid | g | 1.56e-3 ± 1.18e-2 |
| Meal composition | Phosphorus | g | 1.39e-1 ± 1.99e-1 |
| Meal composition | Potassium | g | 3.02e-1 ± 4.02e-1 |

| | | | |
|---|---|---|---|
| Meal composition | Salt | g | 0.65 ± 1.71 |
| Meal composition | Sodium | g | 4.22e-1 ± 7.10e-1 |
| Meal composition | Sugar | g | 12.93 ± 14.55 |
| Meal composition | Vitamin B1 (thiamine) | g | 1.34e-3 ± 1.07e-2 |
| Meal composition | Vitamin B12 (cobalamin) | g | 6.98e-7 ± 3.75e-6 |
| Meal composition | Vitamin B2 (riboflavin) | g | 6.48e-4 ± 7.25e-3 |
| Meal composition | Vitamin B6 | g | 6.65e-4 ± 5.35e-3 |
| Meal composition | Vitamin C | g | 1.28e-2 ± 2.99e-2 |
| Meal composition | Vitamin D | g | 4.01e-7 ± 1.26e-6 |
| Meal composition | Zinc | g | 1.14e-3 ± 2.26e-3 |
| Meal composition | Grains, potatoes and pulses | g | 47.93 ± 88.53 |
| Meal composition | Sweets, salty snacks and alcoholic beverages | g | 33.14 ± 81.02 |
| Meal composition | Non-alcoholic beverages | g | 89.53 ± 143.55 |
| Meal composition | Dairy products, meat, fish, eggs and tofu | g | 53.00 ± 92.35 |
| Meal composition | Vegetables and fruit | g | 51.19 ± 94.99 |
| Meal composition | Oils, fats and nuts | g | 5.91 ± 17.03 |
| Recent dietary intake | Carbohydrate intake in the previous 1 h | g | 9.44 ± 24.88 |
| Recent dietary intake | Carbohydrate intake in the previous 2 h | g | 19.71 ± 34.29 |
| Recent dietary intake | Carbohydrate intake in the previous 3 h | g | 30.92 ± 40.35 |
| Recent dietary intake | Carbohydrate intake in the previous 6 h | g | 60.88 ± 53.57 |
| Recent dietary intake | Fat intake in the previous 1 h | g | 4.54 ± 12.64 |
| Recent dietary intake | Fat intake in the previous 2 h | g | 9.13 ± 17.29 |
| Recent dietary intake | Fat intake in the previous 3 h | g | 13.50 ± 20.18 |
| Recent dietary intake | Fat intake in the previous 6 h | g | 24.97 ± 25.89 |
| Recent dietary intake | Protein intake in the previous 1 h | g | 4.33 ± 12.43 |
| Recent dietary intake | Protein intake in the previous 2 h | g | 8.62 ± 17.17 |
| Recent dietary intake | Protein intake in the previous 3 h | g | 12.44 ± 19.60 |
| Recent dietary intake | Protein intake in the previous 6 h | g | 21.99 ± 23.78 |
| Recent dietary intake | Dietary fiber intake in the previous 1 h | g | 1.25 ± 3.58 |
| Recent dietary intake | Dietary fiber intake in the previous 2 h | g | 2.49 ± 4.88 |
| Recent dietary intake | Dietary fiber intake in the previous 3 h | g | 3.66 ± 5.63 |
| Recent dietary intake | Dietary fiber intake in the previous 6 h | g | 6.77 ± 7.23 |
| Recent dietary intake | Energy intake in the previous 1 h | kcal | 101.43 ± 255.42 |
| Recent dietary intake | Energy intake in the previous 2 h | kcal | 207.69 ± 349.59 |
| Recent dietary intake | Energy intake in the previous 3 h | kcal | 312.60 ± 403.91 |
| Recent dietary intake | Energy intake in the previous 6 h | kcal | 584.53 ± 512.29 |
| Pre-meal glycemic state | Glucose levels at food intake time | mg/dL | 94.23 ± 16.42 |
| Pre-meal glycemic state | Glucose trend over the previous 1 h | mg/dL/h | -4.61 ± 19.43 |
| Pre-meal glycemic state | Glucose trend over the previous 2 h | mg/dL/h | -1.37 ± 11.87 |
| Pre-meal glycemic state | Glucose trend over the previous 4 h | mg/dL/h | -0.09 ± 6.09 |
| Pre-meal glycemic state | Glucose trend over the previous 6 h | mg/dL/h | 0.17 ± 3.95 |
| Meal timing | Time since previous meal | h | 3.37 ± 3.73 |
| Meal timing | Meal time of day | h | 13.36 ± 4.78 |

***Supplementary Table S1.2: Feature description.*** *Description of the features used by the MMER-XGBoost framework. Values represent the mean ± SD across the 50,463 free-living meals, before model scaling and imputation. Observations were available in 50,239, 50,275, 50,303 and 50,320 meals for glucose trends over the preceding 1,2,4, and 6 hours, respectively. The time since the previous meal was available for 49,857 meals. All other features had the complete 50,463 observations.*

| Category | Metric | Unit | Free-living meals (n=50,463): Mean ± SD | Glucose drink (n=1,766): Mean ± SD | White bread (n=1,465): Mean ± SD | White bread with butter (n=1,293): Mean ± SD |
|---|---|---|---|---|---|---|
| MaxGlu | Peak postprandial glucose | mg/dL | 116.49 ± 22.21 | 148.97 ± 28.96 | 133.01 ± 25.54 | 120.20 ± 20.43 |
| PeakDuration | Time above pre-meal glucose | min | 85.40 ± 37.95 | 109.25 ± 17.95 | 110.01 ± 17.34 | 109.17 ± 18.20 |
| Glu120 | Glucose at 120 minutes | mg/dL | 98.99 ± 18.08 | 98.61 ± 25.01 | 103.68 ± 21.93 | 102.62 ± 19.61 |
| iAUC | Positive incremental area under the glucose curve | mg·min/dL | 1,220.05 ± 1,219.97 | 3,470.43 ± 1,773.75 | 2,613.21 ± 1,402.09 | 1,959.21 ± 1,132.07 |
| ΔMaxGlu | Peak glucose increase above baseline | mg/dL | 22.26 ± 18.79 | 62.05 ± 26.47 | 46.22 ± 21.27 | 33.90 ± 16.89 |
| ΔGlu120 | Glucose at 120 minutes minus baseline | mg/dL | 4.76 ± 17.98 | 11.69 ± 23.81 | 16.89 ± 18.93 | 16.33 ± 16.19 |

***Supplementary Table S1.3: PPGR outcome definition.*** *Postprandial glucose response description and distributions in free-living and standardized meals. We report mean ± SD, and sample size per meal type in the column headers.*

| Category | Metric | Unit | Mean ± SD |
|---|---|---|---|
| CGM: glucose distribution | Mean glucose | mg/dL | 95.85 ± 10.49 |
| CGM: glucose distribution | Median glucose | mg/dL | 93.36 ± 9.95 |
| CGM: glucose distribution | Minimum glucose | mg/dL | 55.66 ± 11.15 |
| CGM: glucose distribution | Maximum glucose | mg/dL | 170.62 ± 29.30 |
| CGM: glucose distribution | 25th percentile of glucose | mg/dL | 85.65 ± 9.17 |
| CGM: glucose distribution | 75th percentile of glucose | mg/dL | 103.69 ± 12.20 |
| CGM: glucose variability | Overall glucose SD | mg/dL | 15.47 ± 4.96 |
| CGM: glucose variability | Overall glucose coefficient of variation | % | 16.13 ± 3.78 |
| CGM: glucose variability | Mean daily glucose SD | mg/dL | 13.34 ± 3.60 |
| CGM: glucose variability | Median daily glucose SD | mg/dL | 13.20 ± 3.63 |
| CGM: glucose variability | SD of daily glucose SDs | mg/dL | 3.77 ± 1.91 |
| CGM: glucose variability | Mean daily glucose coefficient of variation | % | 13.94 ± 2.99 |
| CGM: glucose variability | Median daily glucose coefficient of variation | % | 13.78 ± 3.04 |
| CGM: glucose variability | SD of daily glucose coefficients of variation | percentage points | 3.84 ± 1.28 |

| | | | |
|---|---|---|---|
| CGM: glucose variability | Mean amplitude of glycemic excursions | mg/dL | 25.41 ± 11.50 |
| CGM: glucose variability | Mean of daily differences | mg/dL | 13.89 ± 5.11 |
| CGM: glucose variability | Continuous overall net glycemic action over 24 h | mg/dL | 9.58 ± 5.09 |
| CGM: time in range | Total time outside 70-180 mg/dL | min | 1,165.12 ± 2,090.23 |
| CGM: time in range | Total time within 70-180 mg/dL | min | 20,828.39 ± 8,026.60 |
| CGM: time in range | Percentage of readings outside 70-180 mg/dL | % | 5.07 ± 8.27 |
| CGM: glycemic indices | J-index | Index | 12.57 ± 4.11 |
| CGM: glycemic indices | Low blood glucose index | Index | 2.21 ± 1.61 |
| CGM: glycemic indices | High blood glucose index | Index | 0.15 ± 0.65 |
| CGM: glycemic indices | Average daily risk range | Index | 10.00 ± 4.14 |
| CGM: HbA1c estimates | Glucose management indicator | % | 5.60 ± 0.25 |
| CGM: HbA1c estimates | Estimated HbA1c | % | 4.97 ± 0.37 |
| Participant pre-meal glucose summaries | Participant mean pre-meal glucose | mg/dL | 94.12 ± 9.84 |
| Participant pre-meal glucose summaries | Participant SD of pre-meal glucose | mg/dL | 12.82 ± 3.53 |

***Supplementary Table S1.4: Participant-level CGM metrics (N=992).*** *We report mean ± SD. CGM metric calculations excluded the postprandial window following standardized meals. Participant mean and SD of pre-meal glucose were calculated from all free-living and standardized meals.*

#### Supplementary Note 2 | Temporally blocked within-participant cross-validation and leakage control

We employed a subject-wise, temporally contiguous cross-validation strategy that preserves the chronological structure of longitudinal data. For each subject, we partitioned the timeline into five multiday bins of approximately equal duration (“fold”). Each CV iteration held out one fold for validation, and trained on the remaining observations before and/or after it. We excluded training samples within a 6-hour embargo window around the validation fold, because some features use information from up to 6 hours in the past and would otherwise leak across CV iterations.

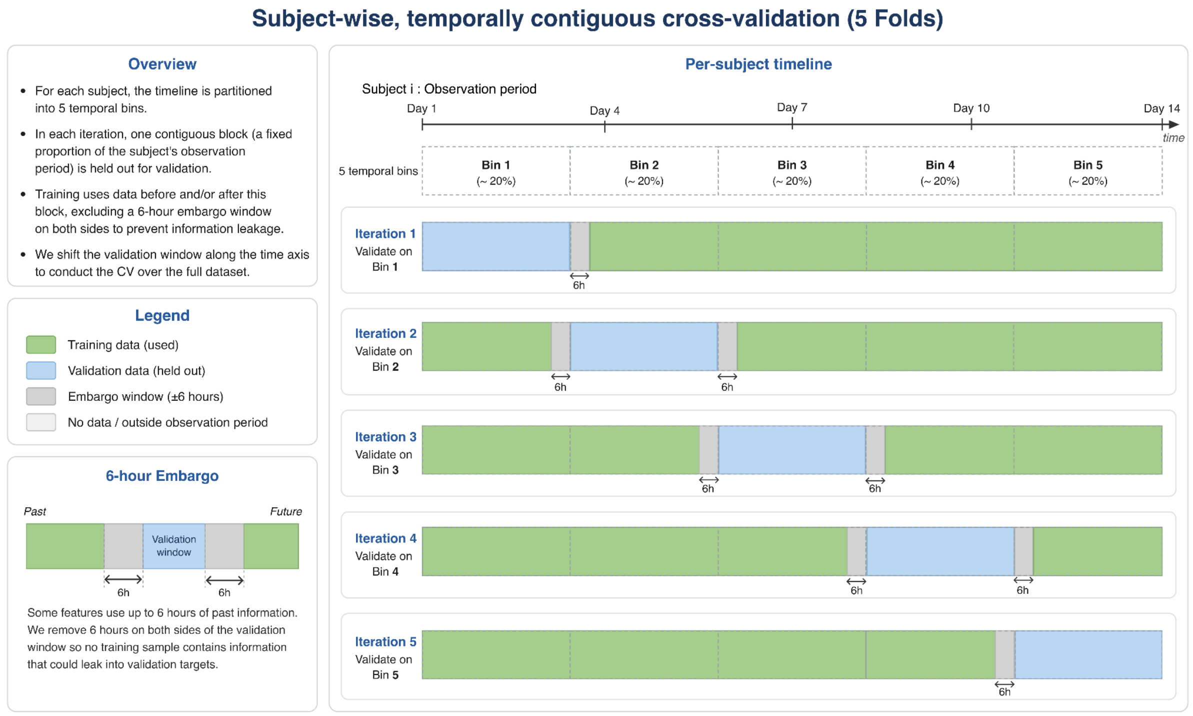


***Supplementary Figure S1:*** *Cross-validation design used for model evaluation.*

#### Supplementary Note 3 | Macronutrient responsiveness after accounting for overall glycemic elevation

We partitioned the covariance matrix of random-effects into four blocks, where i indexes the random intercepts, and s the macronutrient slopes:

$$\tau = \begin{pmatrix} \tau_{ii} & \tau_{is} \\ \tau_{si} & \tau_{ss} \end{pmatrix}$$

Under the model's Gaussian assumptions, we calculated the slope covariance conditional on the intercept as:

$$\tau_{ss\bullet i} = \tau_{ss} - \tau_{si}\,\tau_{ii}^{+}\,\tau_{is}$$

Where + denotes the Moore-Penrose pseudoinverse. This calculation used the fitted covariance matrix from the entire free-living data and population, excluding standardized meals. The statistical uncertainty was assessed using the bootstrap distributions reported below. We eigendecomposed the conditioned covariance matrix $\tau_{ss\bullet i}$ . Principal component signs were chosen to ensure that carbohydrate loadings were positive if a carbohydrate term was among the three largest absolute loadings. Otherwise, the sign was chosen so that the largest absolute loading was positive. We repeated the same operations for each of the 1,000 parametric-bootstrap covariance estimates and calculated 95% percentile confidence intervals. We matched the directions and signs of each bootstrap-derived component loading to the reference components.

The conditioned macronutrient-response subspace was more broadly distributed than the full glycemic phenotype. Its effective dimensionality was 4.31 (95% CI: 4.07-5.56), indicating that macronutrient responsiveness is dominated by at least 4 axes. The first conditioned component explained 37.2% of residual slope-subspace variance (95% CI: 29.1-42.1%), the second explained 24.0% (95% CI: 18.0-26.1%), and the third explained 17.2% (95% CI: 12.0-18.3%). The first two conditioned components explained 61.2% of the residual variance (95% CI: 52.0-63.1%).

| Principal component | Variance explained | Median alignment cosine | Interpretation |
|---|---|---|---|
| $PC_C1$ | 37.2% (29.1-42.1%) | 0.97 | Carbohydrate-versus-fiber attenuation |
| $PC_C2$ | 24.0% (18.0-26.1%) | 0.95 | Protein-versus-fat contrast |
| $PC_C3$ | 17.2% (12.0-18.3%) | 0.95 | Fiber-versus-fat contrast |
| $PC_C4$ | 5.0% (4.9-7.4%) | 0.82 | Smaller residual axis; not interpreted further |

***Supplementary Table S3.1****: Principal component analysis (eigendecomposition) of the conditioned macronutrient-response covariance matrix.*

For iAUC, MaxGlu and Glu120, $PC_C1$ had positive carbohydrate loadings and negative fiber loadings (Supplementary Table S3.2). Across the same outcomes, $PC_C2$ had positive protein loadings and negative fat loadings, whereas $PC_C3$ had positive fiber loadings and negative fat loadings (Supplementary Tables S3.3 and S3.4). Median bootstrap alignment cosines were 0.97, 0.95 and 0.95 for these components, respectively.

| Random effect | PPGR outcome | Loadings |
|---|---|---|
| Carbohydrates | iAUC | +0.54 [+0.47; +0.58] |
| Carbohydrates | MaxGlu | +0.48 [+0.40; +0.51] |
| Carbohydrates | Glu120 | +0.40 [+0.30; +0.48] |
| Fiber | iAUC | -0.34 [-0.39; -0.16] |
| Fiber | MaxGlu | -0.28 [-0.34; -0.15] |
| Fiber | Glu120 | -0.22 [-0.31; -0.10] |
| Protein | iAUC | -0.15 [-0.35; 0.02] |
| Protein | MaxGlu | -0.13 [-0.29; +0.03] |
| Carbohydrates | PeakDuration | +0.13 [+0.12; +0.23] |
| Fiber | PeakDuration | -0.08 [-0.18; -0.01] |

***Supplementary Table S3.2.*** *Ten largest leading loadings of* $PC_C1$ *ordered by absolute magnitude*

| Random effect | PPGR outcome | Loadings |
|---|---|---|
| Protein | iAUC | +0.55 [+0.33; +0.56] |
| Protein | MaxGlu | +0.45 [+0.29; +0.49] |
| Protein | Glu120 | +0.40 [+0.25; +0.44] |
| Fat | iAUC | -0.32 [-0.49; +0.01] |
| Fat | MaxGlu | -0.27 [-0.41; -0.01] |
| Fat | Glu120 | -0.25 [-0.37; +0.00] |
| Fiber | iAUC | -0.17 [-0.44; +0.11] |
| Protein | PeakDuration | +0.15 [+0.11; +0.29] |
| Fiber | MaxGlu | -0.13 [-0.35; +0.05] |
| Fiber | Glu120 | -0.12 [-0.30; +0.05] |

***Supplementary Table S3.3.*** *Ten largest leading loadings of* $PC_C2$ *ordered by absolute magnitude*

| Random effect | PPGR outcome | Loadings |
|---|---|---|
| Fiber | iAUC | +0.47 [+0.25; +0.54] |
| Fat | iAUC | -0.45 [-0.55; -0.18] |
| Fat | Glu120 | -0.34 [-0.41; -0.11] |
| Fat | MaxGlu | -0.32 [-0.46; -0.13] |
| Fiber | MaxGlu | +0.32 [+0.15; +0.42] |
| Fiber | Glu120 | +0.27 [+0.13; +0.37] |
| Fiber | PeakDuration | +0.20 [+0.09; +0.31] |
| Carbohydrates | Glu120 | +0.17 [+0.01; +0.31] |
| Fat | PeakDuration | -0.15 [-0.31; -0.07] |
| Carbohydrates | MaxGlu | +0.15 [+0.02; +0.27] |

***Supplementary Table S3.4.*** *Ten largest leading loadings of* $PC_C3$ *ordered by absolute magnitude*

#### Supplementary Note 4 | Recoverability of participant coordinates as a function of monitoring length

We assessed how quickly an individual's position on the recovered glycemic axes stabilizes as free-living data accumulate. For each monitoring length *d,* from 1 to 14 observed days, we refitted the mixed-effects model using each participant's free-living meals from the first *d* days and extracted the matrix of random effects (BLUPs). Each window model used scaling parameters estimated from its own training data. Before projection, we converted the BLUPs to the reference model's scale. For each reference comparison, we projected the BLUPs onto the eigenvectors of the reference covariance matrix of random effects. Eigenvector signs were chosen so that the largest absolute loading was positive. Agreement was evaluated in the same 853 eligible participants at every monitoring length.

All monitoring-window models and score-agreement analyses were restricted to 853 participants with at least 14 observed days. The 14-day reference was fitted using only their free-living meals from the first 14 observed days. We provided a sensitivity analysis by doing the same comparison to a full-record reference, using all free-living meals from 992 participants, and reported the score agreement in the same 853 participants.

We summarized agreement using the absolute-agreement ICC, ICC(2,1); consistency ICC, ICC(3,1), and Pearson's and Spearman's correlations were reported as supplementary measures. The 14-day reference was fitted to 39,482 free-living meals from the 853 window-eligible participants. The uncapped full-record reference was fitted to 50,463 free-living meals from all 992 prepared participants; score agreement with that reference was evaluated in the same 853 participants. Absolute-agreement ICCs differed by at most 0.038 between references, and both references yielded the same first crossings of 0.75 and 0.90 for each axis.

| **Duration, in days** | **$PC_{Full}$1 scores** | | | | **$PC_{Full}$2 scores** | | | |
|---|---|---|---|---|---|---|---|---|
| | ICC(2,1) | ICC(3,1) | Pearson's r | Spearman's ρ | ICC(2,1) | ICC(3,1) | Pearson's r | Spearman's ρ |
| 1 | 0.51 | 0.51 | 0.51 | 0.47 | 0.25 | 0.25 | 0.26 | 0.24 |
| **2** | 0.64 | 0.64 | 0.64 | 0.62 | 0.32 | 0.32 | 0.33 | 0.27 |
| **3** | 0.75 | 0.75 | 0.76 | 0.74 | 0.38 | 0.38 | 0.41 | 0.34 |
| **4** | 0.82 | 0.82 | 0.83 | 0.80 | 0.49 | 0.49 | 0.52 | 0.45 |
| **5** | 0.86 | 0.86 | 0.87 | 0.85 | 0.56 | 0.56 | 0.61 | 0.55 |
| **6** | 0.89 | 0.89 | 0.90 | 0.88 | 0.66 | 0.66 | 0.69 | 0.64 |
| **7** | 0.91 | 0.91 | 0.92 | 0.91 | 0.72 | 0.72 | 0.74 | 0.71 |
| **8** | 0.94 | 0.93 | 0.94 | 0.93 | 0.79 | 0.79 | 0.80 | 0.77 |
| **9** | 0.95 | 0.95 | 0.96 | 0.95 | 0.83 | 0.83 | 0.84 | 0.82 |
| **10** | 0.97 | 0.97 | 0.97 | 0.97 | 0.88 | 0.88 | 0.88 | 0.86 |

| | | | | | | | | |
|---|---|---|---|---|---|---|---|---|
| **11** | 0.98 | 0.98 | 0.98 | 0.98 | 0.93 | 0.93 | 0.94 | 0.92 |
| **12** | 0.99 | 0.99 | 0.99 | 0.99 | 0.96 | 0.96 | 0.96 | 0.95 |
| **13** | 0.99 | 0.99 | 0.99 | 0.99 | 0.96 | 0.96 | 0.97 | 0.95 |
| **14** | 1.00 | 1.00 | 1.00 | 1.00 | 1.00 | 1.00 | 1.00 | 1.00 |

***Supplementary Table S4.1***: *Recoverability of participant* $PC_{Full}$ *coordinates versus monitoring length (14-day reference).*

| **Duration, in days** | **$PC_{Full}$ 1 scores** | | | | **$PC_{Full}$ 2 scores** | | | |
|---|---|---|---|---|---|---|---|---|
| | ICC(2,1) | ICC(3,1) | Pearson's r | Spearman's ρ | ICC(2,1) | ICC(3,1) | Pearson's r | Spearman's ρ |
| **1** | 0.52 | 0.52 | 0.52 | 0.47 | 0.29 | 0.29 | 0.30 | 0.28 |
| **2** | 0.64 | 0.64 | 0.64 | 0.62 | 0.32 | 0.32 | 0.33 | 0.31 |
| **3** | 0.75 | 0.75 | 0.75 | 0.73 | 0.38 | 0.38 | 0.41 | 0.37 |
| **4** | 0.81 | 0.81 | 0.82 | 0.80 | 0.48 | 0.48 | 0.52 | 0.48 |
| **5** | 0.85 | 0.85 | 0.87 | 0.84 | 0.55 | 0.55 | 0.61 | 0.57 |
| **6** | 0.88 | 0.88 | 0.90 | 0.88 | 0.65 | 0.65 | 0.69 | 0.66 |
| **7** | 0.90 | 0.90 | 0.91 | 0.90 | 0.71 | 0.71 | 0.73 | 0.72 |
| **8** | 0.93 | 0.93 | 0.94 | 0.92 | 0.78 | 0.78 | 0.79 | 0.78 |
| **9** | 0.94 | 0.94 | 0.95 | 0.94 | 0.82 | 0.82 | 0.82 | 0.82 |
| **10** | 0.96 | 0.96 | 0.96 | 0.95 | 0.87 | 0.87 | 0.87 | 0.87 |
| **11** | 0.97 | 0.97 | 0.97 | 0.97 | 0.91 | 0.91 | 0.92 | 0.90 |
| **12** | 0.98 | 0.98 | 0.98 | 0.97 | 0.94 | 0.94 | 0.94 | 0.93 |
| **13** | 0.98 | 0.98 | 0.99 | 0.98 | 0.95 | 0.95 | 0.96 | 0.95 |
| **14** | 0.99 | 0.99 | 0.99 | 0.99 | 0.96 | 0.96 | 0.97 | 0.96 |

***Supplementary Table S4.2***: *Recoverability of participant* $PC_{Full}$ *coordinates versus monitoring length (full-data reference).*

These nested comparisons measure agreement with longer-record estimates. Agreement reaches unity by construction at day 14 only for the 14-day reference.

### Supplementary Note 5 | Associations between glycemic phenotype coordinates and CGM descriptors

We report below the partial Pearson's correlation between CGM metrics and participants' $PC_{Full}$ scores. Associations were adjusted for age, sex, and BMI. Correlation values are given with 95% CI, and the FDR corrected p-value. Corrections were applied once all metrics and both components were tested. Sex was coded as a binary value with 'male'=1.

| **CGM Metrics** | **$PC_{Full}$ 1 scores** | | **$PC_{Full}$ 2 scores** | |
|---|---|---|---|---|
| | Adjusted r [95% CI] | $p_{FDR}$ | Adjusted r [95% CI] | $p_{FDR}$ |
| **Mean Glucose** | 0.63 [0.59, 0.67] | $4.5 \times 10^{-111}$ | -0.54 [-0.58, -0.50] | $6.2 \times 10^{-76}$ |
| **Median Glucose** | 0.57 [0.53, 0.61] | $3.7 \times 10^{-87}$ | -0.59 [-0.63, -0.55] | $2.6 \times 10^{-92}$ |
| **Minimum Glucose** | 0.23 [0.17, 0.29] | $1.1 \times 10^{-13}$ | -0.39 [-0.44, -0.33] | $4.9 \times 10^{-36}$ |
| **Max Glucose** | 0.68 [0.64, 0.71] | $5.0 \times 10^{-133}$ | -0.07 [-0.14, -0.01] | $2.4 \times 10^{-2}$ |
| **Q1 glucose** | 0.52 [0.47, 0.56] | $2.2 \times 10^{-69}$ | -0.61 [-0.65, -0.57] | $5.2 \times 10^{-100}$ |
| **Q3 glucose** | 0.62 [0.58, 0.66] | $2.8 \times 10^{-105}$ | -0.50 [-0.54, -0.45] | $1.2 \times 10^{-62}$ |
| **SD glucose** | 0.52 [0.47, 0.56] | $2.0 \times 10^{-68}$ | 0.05 [-0.01, 0.11] | $1.1 \times 10^{-1}$ |
| **Coefficient of variation of glucose** | 0.38 [0.33, 0.43] | $3.9 \times 10^{-35}$ | 0.35 [0.29, 0.40] | $2.6 \times 10^{-29}$ |
| **Mean of Intraday SD** | 0.67 [0.63, 0.70] | $6.3 \times 10^{-127}$ | 0.06 [-0.00, 0.12] | $7.8 \times 10^{-2}$ |
| **Median of Intraday SD** | 0.65 [0.61, 0.69] | $6.0 \times 10^{-119}$ | 0.04 [-0.03, 0.10] | $2.7 \times 10^{-1}$ |
| **SD of Intraday SD** | 0.42 [0.36, 0.47] | $3.4 \times 10^{-42}$ | 0.10 [0.03, 0.16] | $2.7 \times 10^{-3}$ |
| **Mean of Intraday Coefficient of variation** | 0.47 [0.42, 0.52] | $4.5 \times 10^{-55}$ | 0.37 [0.31, 0.42] | $2.8 \times 10^{-32}$ |
| **Median of Intraday Coefficient of variation** | 0.46 [0.41, 0.51] | $6.1 \times 10^{-53}$ | 0.35 [0.29, 0.40] | $4.7 \times 10^{-29}$ |
| **SD of Intraday Coefficient of variation** | 0.35 [0.29, 0.40] | $1.7 \times 10^{-29}$ | 0.30 [0.24, 0.36] | $6.5 \times 10^{-22}$ |
| **Time Outside Range** | -0.16 [-0.22, -0.10] | $6.2 \times 10^{-7}$ | 0.24 [0.18, 0.30] | $3.4 \times 10^{-14}$ |
| **Time In Range** | -0.03 [-0.09, 0.04] | $4.1 \times 10^{-1}$ | -0.10 [-0.16, -0.04] | $2.5 \times 10^{-3}$ |
| **Percentage Outside Range** | -0.19 [-0.25, -0.13] | $1.2 \times 10^{-9}$ | 0.31 [0.25, 0.37] | $1.5 \times 10^{-23}$ |
| **Mean Amplitude of Glycemic excursions** | 0.23 [0.17, 0.29] | $4.4 \times 10^{-13}$ | -0.25 [-0.31, -0.19] | $2.2 \times 10^{-15}$ |
| **J index** | 0.53 [0.49, 0.58] | $4.5 \times 10^{-73}$ | -0.33 [-0.38, -0.27] | $3.4 \times 10^{-26}$ |

| | | | | |
|---|---|---|---|---|
| **Low Blood Glucose Index** | -0.44 [-0.49, -0.39] | $4.7 \times 10^{-47}$ | 0.49 [0.44, 0.54] | $3.1 \times 10^{-61}$ |
| **High Blood Glucose Index** | 0.29 [0.23, 0.35] | $1.1 \times 10^{-20}$ | -0.18 [-0.24, -0.12] | $1.9 \times 10^{-8}$ |
| **Average Daily Risk Range** | 0.04 [-0.02, 0.10] | $2.3 \times 10^{-1}$ | 0.38 [0.33, 0.44] | $6.3 \times 10^{-36}$ |
| **Mean of Daily Differences** | 0.38 [0.32, 0.43] | $1.1 \times 10^{-34}$ | 0.02 [-0.04, 0.09] | $4.3 \times 10^{-1}$ |
| **Continuous Overall Net Glycemic Action (CONGA) over 24h** | 0.42 [0.37, 0.47] | $4.1 \times 10^{-44}$ | -0.07 [-0.13, -0.01] | $3.7 \times 10^{-2}$ |
| **Glucose Management Indicator** | 0.63 [0.59, 0.67] | $4.5 \times 10^{-111}$ | -0.54 [-0.58, -0.50] | $6.2 \times 10^{-76}$ |
| **Estimated A1C** | 0.63 [0.59, 0.67] | $4.5 \times 10^{-111}$ | -0.54 [-0.58, -0.50] | $6.2 \times 10^{-76}$ |
| **Mean Pre-meal glucose** | 0.63 [0.59, 0.67] | $9.9 \times 10^{-110}$ | -0.57 [-0.61, -0.52] | $6.5 \times 10^{-85}$ |
| **STD Pre-meal glucose** | 0.52 [0.47, 0.56] | $6.8 \times 10^{-69}$ | 0.07 [0.01, 0.13] | $3.2 \times 10^{-2}$ |

***Supplementary Table S5.1. Partial correlations between CGM metrics and glycemic phenotype coordinates.*** *Partial Pearson correlation coefficients (r) and 95% confidence intervals between CGM-derived metrics and PCFull1 and PCFull2 scores, adjusted for age, sex, and BMI. The reported p-values were corrected for multiple testing across all CGM metrics and both phenotype axes using the Benjamini-Hochberg false discovery rate (FDR) procedure.*

#### Supplementary Note 6 | Concordance of glycemic phenotype coordinates and standardized meal challenge responses

We evaluated the relationship between $PC_{Full}1$ and $PC_{Full}2$ scores and responses to the three standardized meal challenges (glucose drink, white bread, and white bread with butter) using three complementary analyses. When a participant consumed the same standardized meal on multiple occasions, their responses were averaged. First, we calculated the unadjusted Pearson correlations between each phenotype coordinate and each of the four PPGR outcomes (MaxGlu, Glu120, iAUC, and PeakDuration) for each challenge type (**Supplementary Table S6.1**). Second, we tested whether each phenotype coordinate explained additional variation in standardized-meal responses beyond age, sex, BMI, mean and coefficient of variation of glucose. For each challenge, outcome and a glycemic axis, we compared a baseline linear model containing these covariates with a full model additionally containing $PC_{Full}1$ or $PC_{Full}2$. We reported the baseline and full adjusted $R^2$, the change in adjusted $R^2$, and p-values from nested F-tests in **Supplementary Table S6.2**. Finally, we compared standardized-meal responses between participants in the lowest and highest quartiles of $PC_{Full}$ scores using two-sided Mann-Whitney U tests (**Supplementary Table S6.3**). We applied Benjamini-Hochberg FDR correction separately within the three families of 24 tests: Pearson's correlation tests, nested-model comparisons, and extreme quartile comparison tests.

| Axis | Outcome | Glucose drinks N=885 | | White bread N=759 | | White bread and Butter N=694 | |
|---|---|---|---|---|---|---|---|
| | | Pearson's r | $p_{FDR}$ | Pearson's r | $p_{FDR}$ | Pearson's r | $p_{FDR}$ |
| $PC_{Full}1$ | MaxGlu | 0.65 | $2.2 \times 10^{-107}$ | 0.66 | $1.8 \times 10^{-95}$ | 0.68 | $7.7 \times 10^{-93}$ |
| $PC_{Full}1$ | Glu120 | 0.46 | $2.8 \times 10^{-47}$ | 0.59 | $3.3 \times 10^{-72}$ | 0.60 | $1.7 \times 10^{-69}$ |
| $PC_{Full}1$ | iAUC | 0.53 | $2.1 \times 10^{-64}$ | 0.51 | $1.5 \times 10^{-50}$ | 0.51 | $2.3 \times 10^{-46}$ |
| $PC_{Full}1$ | PeakDuration | 0.11 | $1.3\times10^{-3}$ | 0.18 | $6.9 \times 10^{-7}$ | 0.15 | $8.4 \times 10^{-5}$ |
| $PC_{Full}2$ | MaxGlu | -0.08 | $2.1 \times 10^{-2}$ | -0.12 | $8.2 \times 10^{-4}$ | -0.16 | $3.4 \times 10^{-5}$ |
| $PC_{Full}2$ | Glu120 | -0.21 | $8.2 \times 10^{-10}$ | -0.22 | $4.2 \times 10^{-9}$ | -0.22 | $1.2 \times 10^{-8}$ |
| $PC_{Full}2$ | iAUC | 0.13 | $2.1 \times 10^{-4}$ | 0.14 | $1.7 \times 10^{-4}$ | 0.14 | $2.7 \times 10^{-4}$ |
| $PC_{Full}2$ | PeakDuration | 0.04 | $2.1 \times 10^{-1}$ | 0.08 | $3.0\times10^{-2}$ | 0.11 | $3.2\times10^{-3}$ |

***Supplementary Table S6.1. Pairwise Pearson correlations between PCFull scores and standardized-challenge outcomes.*** *Values show correlation coefficients (r) and Benjamini-Hochberg FDR-corrected p-values. Sample sizes (N) differ between standardized-meal categories because only participants with usable postprandial responses were included.*

| Standardized meal | PC | PPGR outcome | N | Baseline model Adjusted $R^2$ | Full Model Adjusted $R^2$ | Δ-Adjusted-$R^2$ | F-test $p_{FDR}$ |
|---|---|---|---|---|---|---|---|
| Glucose drink | $PC_{Full}1$ | MaxGlu | 885 | 0.328 | 0.466 | 0.139 | $8.6 \times 10^{-45}$ |
| Glucose drink | $PC_{Full}1$ | Glu120 | 885 | 0.217 | 0.26 | 0.044 | $2.3 \times 10^{-12}$ |
| Glucose drink | $PC_{Full}1$ | iAUC | 885 | 0.164 | 0.334 | 0.17 | $2.0 \times 10^{-44}$ |
| Glucose drink | $PC_{Full}1$ | PeakDuration | 885 | -0.003 | 0.049 | 0.052 | $1.3 \times 10^{-11}$ |
| White bread | $PC_{Full}1$ | MaxGlu | 759 | 0.397 | 0.516 | 0.119 | $2.8 \times 10^{-37}$ |

| | | | | | | | |
|---|---|---|---|---|---|---|---|
| White bread | $PC_{Full}1$ | Glu120 | 759 | 0.412 | 0.476 | 0.064 | $2.6 \times 10^{-20}$ |
| White bread | $PC_{Full}1$ | iAUC | 759 | 0.198 | 0.34 | 0.142 | $2.4 \times 10^{-33}$ |
| White bread | $PC_{Full}1$ | PeakDuration | 759 | 0.004 | 0.038 | 0.035 | $3.2 \times 10^{-7}$ |
| White bread + butter | $PC_{Full}1$ | MaxGlu | 694 | 0.395 | 0.524 | 0.13 | $2.8 \times 10^{-37}$ |
| white bread + butter | $PC_{Full}1$ | Glu120 | 694 | 0.409 | 0.495 | 0.086 | $4.9 \times 10^{-25}$ |
| white bread + butter | $PC_{Full}1$ | iAUC | 694 | 0.19 | 0.334 | 0.144 | $1.0 \times 10^{-30}$ |
| white bread + butter | $PC_{Full}1$ | PeakDuration | 694 | 0.021 | 0.051 | 0.03 | $4.3 \times 10^{-6}$ |
| Glucose drink | $PC_{Full}2$ | MaxGlu | 885 | 0.328 | 0.346 | 0.018 | $1.1 \times 10^{-6}$ |
| Glucose drink | $PC_{Full}2$ | Glu120 | 885 | 0.217 | 0.216 | -0.001 | $7.5 \times 10^{-1}$ |
| Glucose drink | $PC_{Full}2$ | iAUC | 885 | 0.164 | 0.184 | 0.021 | $2.8 \times 10^{-6}$ |
| Glucose drink | $PC_{Full}2$ | PeakDuration | 885 | -0.003 | -0.002 | 0.001 | $1.6 \times 10^{-1}$ |
| White bread | $PC_{Full}2$ | MaxGlu | 759 | 0.397 | 0.415 | 0.018 | $1.9 \times 10^{-6}$ |
| White bread | $PC_{Full}2$ | Glu120 | 759 | 0.412 | 0.419 | 0.007 | $2.5 \times 10^{-3}$ |
| White bread | $PC_{Full}2$ | iAUC | 759 | 0.198 | 0.215 | 0.017 | $4.3 \times 10^{-5}$ |
| White bread | $PC_{Full}2$ | PeakDuration | 759 | 0.004 | 0.013 | 0.009 | $6.0 \times 10^{-3}$ |
| white bread + butter | $PC_{Full}2$ | MaxGlu | 694 | 0.395 | 0.404 | 0.009 | $1.1 \times 10^{-3}$ |
| white bread + butter | $PC_{Full}2$ | Glu120 | 694 | 0.409 | 0.413 | 0.004 | $2.5 \times 10^{-2}$ |
| white bread + butter | $PC_{Full}2$ | iAUC | 694 | 0.19 | 0.201 | 0.01 | $2.2 \times 10^{-3}$ |
| white bread + butter | $PC_{Full}2$ | PeakDuration | 694 | 0.021 | 0.027 | 0.006 | $2.5 \times 10^{-2}$ |

***Supplementary Table S6.2*** *: Nested model tests*

| **Standardized Meal** | **PC** | **PPGR outcome** | **$N_{Q1}$ vs $N_{Q4}$** | **Q1 Median** | **Q4 Median** | **$p_{FDR}$** |
|---|---|---|---|---|---|---|
| Glucose drink | $PC_{Full}1$ | MaxGlu | 222 vs 221 | 127.6 mg/dL | 169.6 mg/dL | $5.3 \times 10^{-51}$ |
| Glucose drink | $PC_{Full}1$ | iAUC | 222 vs 221 | 2333.8 min·mg/dL | 4209.9 min·mg/dL | $1.6 \times 10^{-37}$ |
| Glucose drink | $PC_{Full}1$ | Glu120 | 222 vs 221 | 87.1 mg/dL | 109.3 mg/dL | $8.5 \times 10^{-30}$ |
| Glucose drink | $PC_{Full}1$ | PeakDur | 222 vs 221 | 112.1 min | 116.4 min | $7.5 \times 10^{-4}$ |
| Glucose drink | $PC_{Full}2$ | Glu120 | 222 vs 221 | 101.6 mg/dL | 92.3 mg/dL | $1.1 \times 10^{-5}$ |
| Glucose drink | $PC_{Full}2$ | iAUC | 222 vs 221 | 3212.8 min·mg/dL | 3761.4 min·mg/dL | $3.6 \times 10^{-5}$ |
| Glucose drink | $PC_{Full}2$ | MaxGlu | 222 vs 221 | 153.0 mg/dL | 151.2 mg/dL | $1.9 \times 10^{-1}$ |
| Glucose drink | $PC_{Full}2$ | PeakDur | 222 vs 221 | 112.7 min | 114.0 min | $4.4 \times 10^{-1}$ |
| White bread | $PC_{Full}1$ | MaxGlu | 190 vs 190 | 115.5 mg/dL | 149.4 mg/dL | $1.0 \times 10^{-44}$ |
| White bread | $PC_{Full}1$ | Glu120 | 190 vs 190 | 91.6 mg/dL | 116.1 mg/dL | $6.4 \times 10^{-39}$ |
| White bread | $PC_{Full}1$ | iAUC | 190 vs 190 | 1714.7 min·mg/dL | 3410.0 min·mg/dL | $9.5 \times 10^{-31}$ |
| White bread | $PC_{Full}1$ | PeakDur | 190 vs 190 | 110.6 min | 120 min | $1.0 \times 10^{-9}$ |
| White bread | $PC_{Full}2$ | iAUC | 190 vs 190 | 2554.6 min·mg/dL | 3037.5 min·mg/dL | $3.9 \times 10^{-5}$ |
| White bread | $PC_{Full}2$ | Glu120 | 190 vs 190 | 111.6 mg/dL | 101.6 mg/dL | $1.2 \times 10^{-4}$ |
| White bread | $PC_{Full}2$ | MaxGlu | 190 vs 190 | 138.6 mg/dL | 135 mg/dL | $4.2 \times 10^{-2}$ |
| White bread | $PC_{Full}2$ | PeakDur | 190 vs 190 | 117.2 min | 116.9 min | $5.0 \times 10^{-1}$ |
| White bread and butter | $PC_{Full}1$ | MaxGlu | 174 vs 174 | 104.4 mg/dL | 135 mg/dL | $2.8 \times 10^{-40}$ |
| White bread and butter | $PC_{Full}1$ | Glu120 | 174 vs 174 | 90.3 mg/dL | 114.0 mg/dL | $2.7 \times 10^{-35}$ |
| White bread and butter | $PC_{Full}1$ | iAUC | 174 vs 174 | 1229.1 min·mg/dL | 2460.4 min·mg/dL | $8.2 \times 10^{-24}$ |
| White bread and butter | $PC_{Full}1$ | PeakDur | 174 vs 174 | 110.3 min | 119.6 min | $1.5 \times 10^{-4}$ |
| White bread and butter | $PC_{Full}2$ | Glu120 | 174 vs 174 | 107.3 mg/dL | 99.1 mg/dL | $1.4 \times 10^{-5}$ |

| | | | | | | |
|---|---|---|---|---|---|---|
| White bread and butter | $PC_{Full}2$ | iAUC | 174 vs 174 | 1682.3 min·mg/dL | 2141.4 min·mg/dL | $2.9 \times 10^{-5}$ |
| White bread and butter | $PC_{Full}2$ | PeakDur | 174 vs 174 | 112 min | 120 min | $6.9 \times 10^{-4}$ |
| White bread and butter | $PC_{Full}2$ | MaxGlu | 174 vs 174 | 124.2 mg/dL | 119.6 mg/dL | $8.4 \times 10^{-3}$ |

**Supplementary Table S6.3** : Extreme-quartile Mann-Whitney comparisons of PPGR outcomes across standardized meals.

#### Supplementary Note 7 | Cross-fitted reproducibility of glycemic phenotype coordinates in held-out participants

We assessed the out-of-sample reproducibility using five-fold cross-fitting at the population-level. Participants were assigned to five mutually exclusive folds, with all meals from a given participant retained in the same fold. We excluded standardized meals from both model fitting and from $PC_{Full}$ score estimations. At each CV iteration, the MMER-XGBoost model was fitted on four folds and applied to the remaining held-out fold. Held-out participants did not contribute to the estimation of the non-linear population-response surface, or the participant-level random-effect covariance. We estimated feature and outcome scaling parameters from the training participants in each fold. The same training-derived scalers were applied to the held-out participants.

We eigendecomposed the 20×20 random-effects covariance matrix estimated  in each training fold to define the phenotype axes. For each group of held-out participants, we split meals into halves based on chronological and alternating odd- and even-indexed meal-splitting schemes. Both schemes produced disjoint meal sets, with chronological halves separating earlier and later periods, and alternating subsets spanning the same observation period.

We separately estimated the participant-specific random effects from subsets A and B using the fitted training-fold models. The fixed-effect surface, random-effect covariance and scaling parameters were not re-estimated in the held-out fold. Each subset therefore yielded a separately estimated BLUP vector for the same participant under an identical model specification. The two BLUP vectors were projected onto the corresponding training-fold eigenvectors. We divided the estimated scores by the square root of the associated eigenvalue to place coordinates from different folds on a common standard-deviation scale.

We summarized the agreement between the paired estimates using the absolute agreement intraclass correlation ICC(2,1), with 95% confidence intervals. We quantified the consistency using the ICC(3,1) to identify any systematic shift between the two meal subsets.

| Analysis | Odd vs even meals | | Early vs late meals | |
|---|---|---|---|---|
| | ICC(2,1) [95% CI] | ICC(3,1) [95% CI] | ICC(2,1) [95% CI] | ICC(3,1) [95% CI] |
| Pooled $PC_{Full}$ 1 | 0.81 [0.79, 0.83] | 0.81 [0.79, 0.83] | 0.74 [0.71, 0.77] | 0.74 [0.71, 0.77] |
| Pooled $PC_{Full}$ 2 | 0.52 [0.48, 0.57] | 0.52 [0.48, 0.57] | 0.48 [0.43, 0.53] | 0.48 [0.44, 0.53] |

***Supplementary Table S7.1: Pooled ICCs for cross-fitted PC1 and PC2 scores under two split-half designs.*** *Absolute agreement, ICC(2,1), and consistency, ICC(3,1), with 95% confidence intervals.*

| Analysis | PC axis | N | ICC(2,1) [95% CI] | ICC(3,1) [95% CI] |
|---|---|---|---|---|
| Fold 1 | $PC_{Full}$ 1 | 199 | 0.84 [0.79, 0.87] | 0.84 [0.79, 0.87] |
| Fold 1 | $PC_{Full}$ 2 | 199 | 0.48 [0.36, 0.58] | 0.48 [0.36, 0.58] |
| Fold 2 | $PC_{Full}$ 1 | 199 | 0.81 [0.75, 0.85] | 0.81 [0.75, 0.85] |
| Fold 2 | $PC_{Full}$ 2 | 199 | 0.58 [0.48, 0.67] | 0.58 [0.48, 0.67] |
| Fold 3 | $PC_{Full}$ 1 | 198 | 0.83 [0.78, 0.87] | 0.83 [0.78, 0.87] |
| Fold 3 | $PC_{Full}$ 2 | 198 | 0.49 [0.37, 0.59] | 0.49 [0.37, 0.58] |
| Fold 4 | $PC_{Full}$ 1 | 198 | 0.80 [0.74, 0.85] | 0.80 [0.74, 0.85] |

| | | | | |
|---|---|---|---|---|
| Fold 4 | $PC_{Full}$ 2 | 198 | 0.53 [0.42, 0.62] | 0.53 [0.42, 0.62] |
| Fold 5 | $PC_{Full}$ 1 | 198 | 0.80 [0.74, 0.85] | 0.80 [0.74, 0.85] |
| Fold 5 | $PC_{Full}$ 2 | 198 | 0.50 [0.39, 0.60] | 0.50 [0.39, 0.60] |

***Supplementary Table S7.2:*** *Fold-specific absolute-agreement and consistency ICC for cross-fitted PC1 and PC2 scores estimated from alternating split (odd- and even-indexed) meal subsets.*

| **Analysis** | **PC axis** | **N** | **ICC(2,1) [95% CI]** | **ICC(3,1) [95% CI]** |
|---|---|---|---|---|
| Fold 1 | $PC_{Full}$ 1 | 199 | 0.73 [0.66, 0.79] | 0.73 [0.66, 0.79] |
| Fold 1 | $PC_{Full}$ 2 | 199 | 0.27 [0.14, 0.40] | 0.28 [0.15, 0.40] |
| Fold 2 | $PC_{Full}$ 1 | 199 | 0.72 [0.64, 0.78] | 0.72 [0.64, 0.78] |
| Fold 2 | $PC_{Full}$ 2 | 199 | 0.61 [0.51, 0.69] | 0.61 [0.51, 0.69] |
| Fold 3 | $PC_{Full}$ 1 | 198 | 0.78 [0.72, 0.83] | 0.78 [0.72, 0.83] |
| Fold 3 | $PC_{Full}$ 2 | 198 | 0.46 [0.34, 0.56] | 0.46 [0.34, 0.56] |
| Fold 4 | $PC_{Full}$ 1 | 198 | 0.73 [0.66, 0.79] | 0.73 [0.65, 0.79] |
| Fold 4 | $PC_{Full}$ 2 | 198 | 0.52 [0.41, 0.61] | 0.52 [0.41, 0.61] |
| Fold 5 | $PC_{Full}$ 1 | 198 | 0.73 [0.66, 0.79] | 0.73 [0.66, 0.79] |
| Fold 5 | $PC_{Full}$ 2 | 198 | 0.47 [0.35, 0.57] | 0.47 [0.35, 0.57] |

***Supplementary Table S7.3:*** *Fold-specific absolute-agreement and consistency ICC for cross-fitted PC1 and PC2 scores estimated from temporal split meal subsets.*

### Supplementary Note 8 | Robustness of the glycemic phenotype to baseline-relative outcome definitions

We repeated the analyses after expressing MaxGlu and Glu120 relative to premeal glucose (respectively named ΔMaxGlu and ΔGlu120). The model comparison remained mostly unchanged: MMER-XGBoost was the best-performing model, with a multivariate $R^2$ of 0.48 across all meals, and $R^2$ of 0.24 for predictions of standardized meal responses (see **Supplementary Table S8.1**). The estimated between-participant structure was also highly preserved. The effective dimensionality was 2.13 compared with 2.00 in the primary analysis. $PC_{Full}1$ and $PC_{Full}2$ explained 67.1% and 11.0% of variance, respectively, compared with 69.3% and 11.8% in the primary analyses. Their loading vectors were nearly identical between specifications (cosine similarity = 0.996 and 0.992), and participant scores were strongly concordant (Pearson's r = 0.998 and 0.975 for $PC_{Full}1$ and $PC_{Full}2$, respectively). The conditioned macronutrient-response structure was similarly preserved (loading cosine = 0.992 for $PC_C1$ and 0.993 for $PC_C2$).

| **Models** | **Free-living and standardized meals** N= 54,987 | | | | | **Standardized meals only** N= 4524 | | | | |
|---|---|---|---|---|---|---|---|---|---|---|
| | **Multivariate** $R^2$ | **Outcome-specific $R^2$ (r)** | | | | **Multivariate** $R^2$ | **Outcome-specific $R^2$ (r)** | | | |
| | | *ΔMaxGlu* | *Peak Duration* | *ΔGlu120* | *iAUC* | | *ΔMaxGlu* | *Peak Duration* | *ΔGlu120* | *iAUC* |
| **MMER-XGBoost** Random intercept and slopes | 0.48 | 0.52 (0.72) | 0.44 (0.67) | 0.46 (0.68) | 0.50 (0.70) | 0.24 | 0.36 (0.63) | 0.13 (0.40) | 0.17 (0.41) | 0.32 (0.59) |
| **MMER-XGBoost** Random intercept only | 0.47 | 0.51 (0.72) | 0.44 (0.67) | 0.46 (0.68) | 0.49 (0.70) | 0.21 | 0.31 (0.60) | 0.14 (0.41) | 0.13 (0.37) | 0.26 (0.55) |
| **XGBoost** No random effects | 0.42 | 0.44 (0.66) | 0.43 (0.65) | 0.41 (0.64) | 0.42 (0.65) | 0.09 | 0.13 (0.43) | 0.11 (0.35) | 0.03 (0.20) | 0.09 (0.36) |
| **LMMER** Random intercept and slopes | 0.37 | 0.36 (0.60) | 0.34 (0.58) | 0.41 (0.64) | 0.36 (0.60) | -0.10 | -0.24 (0.43) | -0.19 (0.35) | 0.16 (0.40) | -0.12 (0.45) |
| **LMMER** Random intercept only | 0.35 | 0.34 (0.59) | 0.33 (0.58) | 0.40 (0.63) | 0.34 (0.59) | -0.16 | -0.32 (0.39) | -0.23 (0.35) | 0.12 (0.36) | -0.20 (0.41) |
| **Linear Regression** No random effects | 0.28 | 0.25 (0.50) | 0.30 (0.55) | 0.34 (0.58) | 0.25 (0.50) | -0.31 | -0.51 (0.10) | -0.35 (0.29) | 0.01 (0.20) | -0.40 (0.11) |

**Supplementary Table S8.1**. **Model comparison using baseline-relative outcomes.**

| Axis | Original variance explained | Baseline-relative outcomes Variance explained | Loading cosine similarity | Pearson's r correlation between scores |
|---|---|---|---|---|
| First axis | 69.3% | 67.1% | 0.996 | **0.998** |
| Second axis | 11.8% | 11.0% | 0.992 | **0.975** |

***Supplementary Table S8.2. Stability of glycemic phenotype geometry using baseline-relative outcome definitions.*** *Comparison of the original PPGR axes with an alternative specification in which MaxGlu and Glu120 were expressed relative to pre-meal glucose (ΔMaxGlu and ΔGlu120). We matched leading components across specifications and compared their variance explained, cosine-similarity between corresponding loading-vectors, and concordance of participant scores.*
